# Multi-ancestry GWAS of Long COVID identifies immune-related loci and etiological links to chronic fatigue syndrome, fibromyalgia and depression

**DOI:** 10.1101/2024.10.07.24315052

**Authors:** Ninad S. Chaudhary, Catherine H. Weldon, Priyanka Nandakumar, 23andMe Research Team, Michael V. Holmes, Stella Aslibekyan

## Abstract

The etiology of Long COVID is poorly understood despite its estimated global burden of 65 million cases. There exists a paucity of genetic studies that can shed light on potential mechanisms leading to Long COVID. Using consented and genotyped data from 23andMe adult research participants, we conducted the largest multi-ancestry meta-analysis of genome-wide association studies of Long COVID across European (42,899 cases, 94,721 controls), Latinx (8,631 cases, 20,351 controls), and African-American (2,234 cases, 5,596 controls) genetic ancestry groups. GWAS of Long COVID identified three genome-wide significant loci (*HLA-DQA1–HLA-DQB, ABO, BPTF–KPAN2*–*C17orf58*). Functional analysis of these genes points to underlying immune and thrombo-inflammatory mechanisms. We present evidence of shared genetic architecture (genetic correlation p-value < 0.001) of Long COVID with thirteen phenotypes of similar symptomatology or pathophysiology. We identified potential causal roles from liability to chronic fatigue (Mendelian randomization OR=1.59, 95% CI[1.51,1.66]), fibromyalgia (OR=1.54, 95% CI[1.49,1.60]), and depression (OR=1.53, 95% CI[1.46,1.61]) with Long COVID, which replicated in the COVID-19 Host Genetics Initiative data, and which are unlikely to originate from collider bias. These findings can help identify populations vulnerable to Long COVID and inform future therapeutic approaches.

## INTRODUCTION

Long COVID (post-acute sequelae of SARS-CoV2-infection or post-acute COVID-19 syndrome) is a debilitating condition characterized by symptoms persisting for months following the acute infection. Since the global spread of SARS-CoV2, more than 65 million people have reported experiencing symptoms that can be classified as Long COVID (Davis et al. 2023). It is estimated that 7.3% of SARS-CoV2 positive individuals and 52% of those hospitalized due to COVID-19 in the US (Robertson et al. 2023; Taquet et al. 2021; O’Mahoney et al. 2023) subsequently develop Long COVID. Individuals experiencing Long COVID predominantly report fatigue, cognitive impairment and breathlessness impacting daily activities (O’Mahoney et al. 2023; Ford et al. 2024; Lopez-Leon et al. 2021).

Vaccination efforts have been shown to reduce the risk of developing Long COVID symptoms in SARS-CoV2 positive individuals (Català et al. 2024; Robertson et al. 2023). However, among individuals with Long COVID, it is not clear whether vaccination attenuates symptoms, making it imperative to continue identifying treatment options for Long COVID (Notarte et al. 2022; Tran et al. 2023). NIH-funded clinical trials are underway for exploring the efficacy of Paxlovid for the treatment of Long COVID. This represents just one potential therapeutic strategy: lack of knowledge of the underlying etiological mechanisms represents an ongoing challenge to developing evidence-based treatment options.

Genome-wide association studies (GWAS) of acute COVID-19 susceptibility and severity have contributed in establishing the role of immune mechanisms in response to acute SARS-CoV2 infection (Shelton et al. 2021; COVID-19 Host Genetics Initiative 2023). In contrast, the genetic evidence identifying biological mechanisms in Long COVID is limited (Ledford 2024). Mechanisms that can lead to Long COVID may include persistence of viral burden, autoimmune response, and inflammation. A recent GWAS of Long COVID by the COVID-19 Host Genetics Initiative (HGI) identified a single risk locus (*FOXP4* gene) for Long COVID at chromosome 6 (Lammi et al. 2023). This locus has previously been linked to acute COVID-19 susceptibility and severity (COVID-19 Host Genetics Initiative 2023), recognised risk factors for Long COVID (Whitaker et al. 2022). However, the mechanisms underlying the *FOXP4* genetic signal, postulated to include lung function, explain only a part of the multi-systemic response that is postulated to occur in Long COVID. Overall, the mechanistic understanding of Long COVID continues to be elusive. Larger-scale GWAS of Long COVID restricted to previously COVID-19 infected individuals are well suited to identifying genetic loci conferring risk of Long COVID, while controlling for susceptibility, and may help inform treatment strategies.

Epidemiological studies have also identified similarities between phenotypic clusters of Long COVID and other post-viral syndromes and chronic conditions. (Sherif et al. 2023; Davis et al. 2023; Thompson et al. 2023). This posits the question of whether a pre-existing chronic condition can increase the risk of developing Long COVID. By leveraging GWAS data on these conditions, we can further elucidate disease etiology to identify putative causal risk factors that may shed light on mechanistic pathways. The 23andMe research cohort provides an opportunity to investigate the genetic architecture of Long COVID and to conduct post-GWAS translational studies such as Mendelian randomization to shed light on potential etiology. Accordingly, using ancestrally-diverse data from the 23andMe database, we conducted GWAS in three ancestries and performed meta-analysis to identify genetic variants associated with Long COVID in a cohort of COVID-19 infected individuals. We further investigated genetic correlations between Long COVID and symptomatologically similar phenotypes and explored their potential causal relevance, including the role of acute COVID-19 severity, through Mendelian randomization.

## METHODS

### Study population and data collection

Details of the 23andMe Research Cohort have been previously described (Shelton et al. 2021). Self-reported data were obtained from online survey questionnaires on demographics, socio-behavioral characteristics, and health conditions. Data on Long COVID were collected through a survey designed to inform about the symptomatic burden and impact on daily living activities. Participants who had previously reported to have COVID-19 diagnosis or tested positive for SARS-CoV2 were invited via email to participate in the Long COVID survey. Additional respondent characteristics, such as age, sex, pre-existing conditions (high blood pressure, diabetes, chronic kidney disease, coronary heart disease, cardiac arrhythmias, high cholesterol, history of blood clots, and chronic obstructive pulmonary disease), educational attainment, smoking status were queried in the baseline COVID-19 surveys if the data were missing from previously distributed health surveys. Data collected between August 2021 and February 2024 were used for the current work (**Figure 1**).

**Figure 1:**
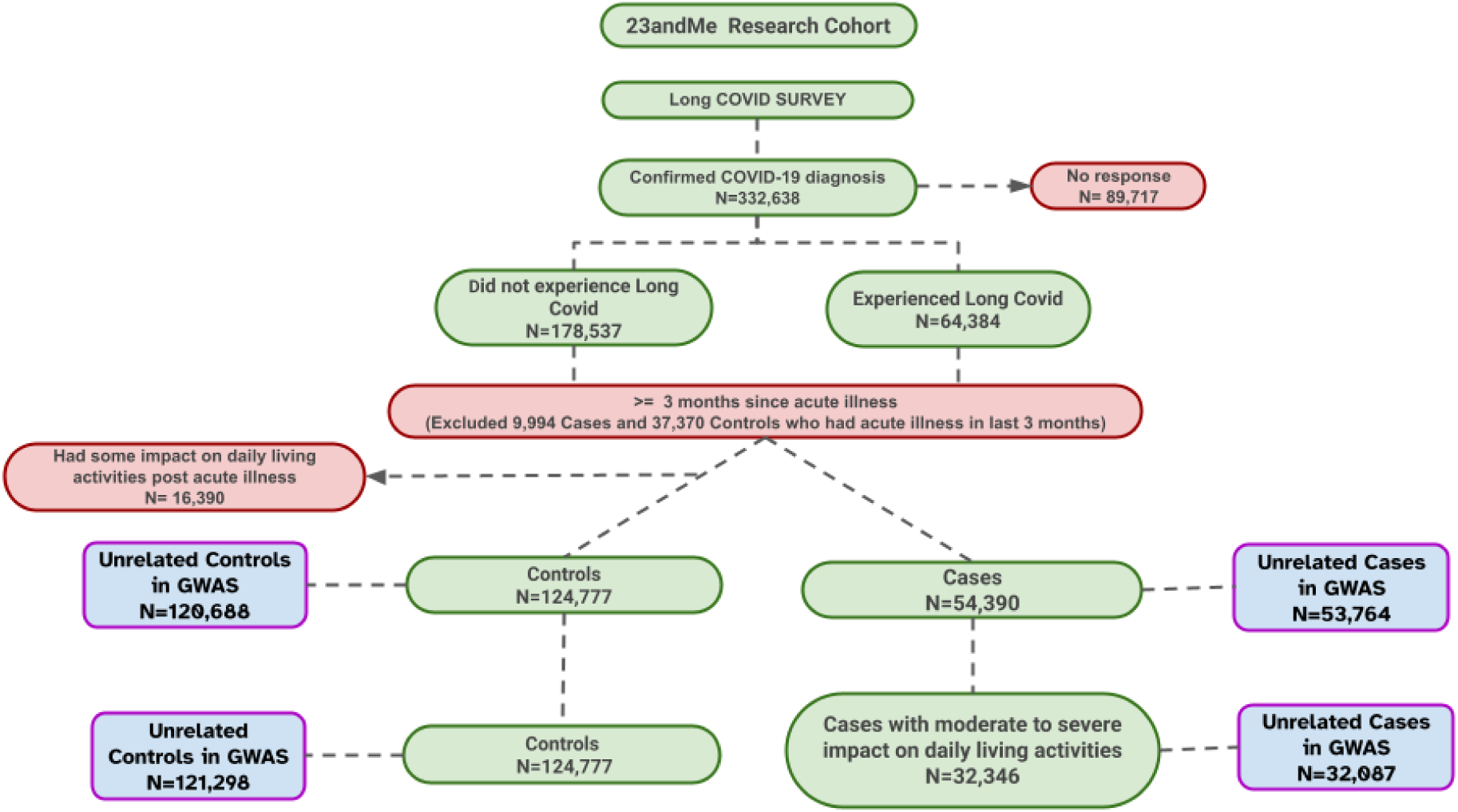
Flowchart of 23andMe participants who participated in Long COVID survey and were included in the analysis. 47,364 participants were excluded if the symptoms are reported less than 3 months from acute illness; 16,390 participants were excluded from the control group if they reported some impact (selected option other than “not at all”) on daily living activities post acute illness. Participants of other ancestries (East Asian, South Asian, and Middle Eastern) present in 23andMe research database were not included in the base cohort due to insufficient sample size.

### Data availability

The full GWAS summary statistics for the 23andMe discovery data set will be made available through 23andMe to qualified researchers under an agreement with 23andMe that protects the privacy of the 23andMe participants. Datasets will be made available at no cost for academic use. Please visit https://research.23andme.com/collaborate/#dataset-access/ for more information and to apply to access the data.

### Ethics statement

Study participants provided informed consent. The recruitment, informed consent, and collection of online data via surveys were conducted according to our human participants research protocol, which was reviewed and approved by Ethical and Independent Review Services, a private institutional review board (http://www.eandirreview.com). As of 2022, E&I Review Services is part of Salus IRB (https://www.versiticlinicaltrials.org/salusirb).

### Phenotype definition

In contrast to prior Long COVID GWAS using population controls (Lammi et al. 2023), only participants who were diagnosed with COVID-19 or had tested positive for COVID-19 were included in the main analysis. We first excluded those who had reported to have had acute COVID-19 illness within the last three months for consistency with the widely accepted definition whereby to be eligible to be a Long COVID case, it occurred three months after the acute infection (Soriano et al. 2022). Long COVID was defined using self-reported data on the question: “*Have you* experienced *post-acute COVID-19 syndrome (also called ’Long COVID’ or ’Long Haulers’)?*”. Those who responded “yes” to the question were defined as cases of Long COVID. Participants who responded “No” to the question were defined as controls.

The data on impact on daily life activities were obtained among individuals who reported having a COVID-19 diagnosis and symptoms lasting for one month or longer since illness from the question “On average, how much has your persistent COVID-19 symptoms impacted your normal activities? If you have fully recovered, please respond based on your experience before you fully recovered”. Participants’ responses were recorded on a 5 point Likert scale of “not at all, slightly, moderately, quite a bit, extremely”. Among controls, 16,390 individuals had responded to having some impact on daily activities post acute illness and were excluded from this group. In a sensitivity analysis, we modified the definition for Long COVID cases incorporating the impact of Long COVID symptoms on daily life (referred to as “Long COVID Impact” phenotype) (Lammi et al. 2023; Soriano et al. 2022). Long COVID Impact phenotype was defined as those who self-reported Long COVID, and had moderate to severe impact of Long COVID symptoms on daily living activities. The impact was considered moderate to severe if participants chose the following answer options: moderately, quite a bit, or extremely (N=32,346).

### Genotyping

DNA extraction and genotyping were performed on saliva samples by Clinical Laboratory Improvement Amendments and Collage of American Pathologists-accredited clinical laboratories of Laboratory Corporation of America. The details of genotyping platforms and imputation quality controls have been previously described (Chaudhary et al. 2024). Approximately 110 million variants were available for genome-wide analyses after the quality control procedures. Genotyped data were further analyzed to determine the ancestry of the participants using a genetic ancestry classification algorithm.

Subsequent analyses were performed separately for each ancestry. To limit our analyses to unrelated individuals, we selected participants such that no two individuals shared more than 700cM of DNA identical by descent. In a scenario where a case and a control have at least 700cM of DNA identical by descent, we removed the control from the analytical sample. After excluding unrelated individuals while prioritizing the retention of cases, the analytical cohort for GWAS consisted of 53,764 cases and 120,688 controls for Long COVID, and 32,087 cases and 121,298 controls for Long COVID Impact.

### Imputation of classical HLA Alleles

Imputation of *HLA* alleles was performed using the HIBAG13 R package (Zheng et al. 2014). HIBAG is an attribute bagging based statistical method that uses a pre-fit classifier to impute the allelic dosages of *HLA* alleles. This classifier was trained from a large internal database of individuals with known *HLA* alleles and SNP variation within the HLA region. We used the default setting of HIBAG and used a multi-population reference panel from 100,000 samples of 23andMe internal data and other external data. Approximately 98% of the tagging SNPs used in HIBAG were genotyped and passed quality control on 23andMe’s platform. We imputed allelic dosage for *HLA-A, B, C, DPA1, DPB1, DQA1, DQB1* and *DRB1* loci at the p-group resolution where *HLA* alleles that encode the same protein sequence for the peptide binding domains (exon 2 and 3 for class I and exon 2 only for class 2) are grouped together and designated with 4 digits (Tian et al. 2017). Similar to SNP imputation, we measured imputation quality using r^2^ which ranged from 0.72 to 1.00. Overall, there were 855 imputed classical HLA alleles at four-digit resolution available for analysis.

### Ancestry determination

Ancestries in the 23andMe database are determined using a classifier algorithm based on analysis of local ancestry (Durand et al. 2014). Phased genotyped data were first partitioned into windows of about 300 SNPs and a support vector machine (SVM) approach was applied within each window to classify individual haplotypes into one of 45 worldwide reference populations. The SVM classifications are then fed into a hidden Markov model (HMM) that accounts for switch errors and incorrect assignments, and gives probabilities for each reference population in each window. Finally, we used simulated admixed individuals to recalibrate the HMM probabilities so that the reported assignments are consistent with the simulated admixture proportions. We aggregated the probabilities of the 45 reference populations into six main ancestry categories (European, African-American, Latinx, East Asian, South Asian, Middle Eastern) using a predetermined threshold (Durand et al. 2014). African Americans and Latinx were admixed with broadly varying contributions from Europe, Africa and the Americas. No single threshold of genome-wide ancestry could effectively discriminate between African Americans and Latinx. However, the distributions of the length of segments of European, African and American ancestry are very different between African Americans and Latinx, because of distinct admixture timing between the three ancestral populations in the two ethnic groups. Therefore, we trained a logistic classifier that took the participant’s length histogram of segments of African, European and American ancestry, and predicted whether the customer is likely African American or Latinx.

## STATISTICAL ANALYSIS

### Multi-ancestral GWAS

Within each ancestral group, we performed logistic regression for Long COVID with additive allelic effects for each variant. The model was adjusted for age, age-squared, sex, and the top five within-ancestry principal components to account for residual population structure and genotyping platform. The genome-wide threshold for significant association for each variant was < 5.0 X 10^-8^. The summary statistics were corrected for inflation using genomic control when the inflation factor was greater than one and presented using Manhattan plots. We then constructed regions of significant association by identifying all SNPs with P < 10^-5^ within the vicinity of a genome-wide significant association. We defined a distance between neighboring regions of association of at least 250kb to identify a significant locus. The SNP with the smallest P-value within each region was identified as the index variant. We also calculated a credible set of variants within each region based on Bayesian approach to refine the association signals (Wellcome Trust Case Control Consortium et al. 2012). The regional LD plots for each index variant were constructed. The analyses were conducted separately in each genetic ancestry cohort (Europeans, African-Americans, and Latinx). Participants classified as other ancestral groups (South Asian, East Asian, and Middle Eastern) were excluded due to insufficient GWAS sample size. We also conducted logistic regression between *HLA* loci and Long COVID adjusting for above mentioned covariates. Previous studies have shown that *HLA* imputed alleles are associated with viral infections as individuals carrying the same *HLA* allelic dosage recognize the same viral epitope (Tian et al. 2017; Augusto et al. 2023; Debebe et al. 2020). We accordingly determined the associations between *HLA* alleles imputed till four-digit resolution and Long COVID by ancestry in the region of statistically significant *HLA* loci.

### Meta-Analysis

We then performed a multi-ancestry meta-analysis of above GWAS using the inverse variance fixed effect model. Only variants with minor allele frequency of more than one percent in all ancestries were considered for meta-analysis. The associations were further adjusted for inflation and tested for heterogeneity between genetic ancestry groups using the Cochran’s Q test. The effect estimates of index variants on meta-analyses are presented as odds ratios (OR) on an additive allelic scale.

### Functional annotation of GWAS index variants

To perform variant-to-gene mapping, hypotheses of functionally relevant genes are generated by annotating the index variants. Functional variants for mapping include coding variants (annotated by the Ensembl Variant Effect Predictor (VEP) v109 44), eQTLs, and pQTLs. The eQTL annotation resources consist of a comprehensive collection of standardized variants impacting gene expression in various tissues obtained from publicly available datasets (GTEx Consortium 2020; Qiu et al. 2018; Kettunen et al. 2012; Franzén et al. 2016; Raj et al. 2014; Kerimov et al. 2021; Lappalainen et al. 2013; Koolpe et al. 1985; Craig et al. 2021) and datasets processed by the 23andMe eQTL pipeline. The pQTL annotation resources similarly include a collection of curated protein QTLs from relevant public datasets (Zhang et al. 2022; Ferkingstad et al. 2021).

eQTL calling was performed with one of two versions of 23andMe pipelines, depending on the dataset in question (**Supplementary File**). The first pipeline used FastQTL (Ongen et al. 2016) in permutation mode, restricting all tests to variants within a window defined to be 1Mbp up- or downstream of a given gene’s transcription start site (TSS) (**Supplementary File Table 1**). Variants tested were single nucleotide polymorphisms with an in-sample MAF ≥ 1%, to avoid errors in detection or mapping of larger genetic variants in cross-ancestry comparisons, and models were adjusted for age (if available), sex, probabilistic estimation of expression residuals (PEER) factors (Stegle et al. 2012), genetic PCs, and per-dataset covariates. For each gene, the index variant was identified by the minimal permutation p-value. eQTLs were called then on lead variants if they passed a 5% false discovery rate (FDR) filter using Storey’s q-value (Storey 2002) methodology. Conditional eQTLs were identified via FastQTL’s permutation mode, by using each eQTL as an additional covariate in the model for a given gene. The lead conditional eQTL for all genes were again FDR controlled at 5%, and a maximum of 10 conditional steps were run. Finally, for a set of conditional eQTLs for a given gene, a joint model was fit, and the final eQTL callset consisted of those eQTLs whose joint model test passed a 5% FDR filter. eQTLs were only called for genes classified as one of ’protein_coding’, ’miRNA’, ’IG_C_gene’, ’IG_D_gene’, ’IG_J_gene’, ’IG_V_gene’,’TR_C_gene’, ’TR_D_gene’, ’TR_J_gene’, ’TR_V_gene’ as defined in GENCODE (Harrow et al. 2012).

The second version of the 23andMe pipeline uses strand-aware RNA-seq quantification, and the eQTLs were called using SusieR package (Wang et al. 2020) instead of FastQTL, with expression PCs (selected with the elbow method) replacing PEER factors in the modeling, and using the GENCODE v43 gene model (**Supplementary File Table 2**). The pipeline natively generated credible sets with a set probability to contain a SNP tagging the causal variant.

### Genetic correlation of Long COVID with phenotypically similar traits

Though the exact biological mechanisms of Long COVID remain to be determined, it is proposed that Long COVID is a consequence of the multi-system effect of acute-phase COVID-19 infection (Thompson et al. 2023). The phenotypic clusters and pathophysiology of Long COVID overlap with post-viral syndromes, dystonia, chronic fatigue/MFS, endothelial dysfunction, and conditions associated with immune dysregulation and coagulopathies. To understand whether these phenotypic and pathophysiological similarities have genetic underpinnings, we obtained the genetic correlations between Long COVID and symptomatologically similar phenotypes in participants of European ancestry using LD score regression (Bulik-Sullivan et al. 2015).

We selected phenotypes if the phenotypes were available in the 23andMe database and 1) the symptomatology of the phenotype overlapped with the symptoms of Long COVID (chronic pain, chronic obstructive pulmonary disease, polymyalgia rheumatica, chronic fatigue/myalgic encephalitis, fibromyalgia, Alzheimer’s disease, polymyositis, hypersensitivity pneumonitis, lupus, complex regional pain syndrome, asthma); and/or 2) the phenotype had known etiological precedent of viral illness (EBV-related illness such as chronic fatigue syndrome/myalgic encephalitis, dysautonomia, multiple sclerosis, Guillain-Barre syndrome, chronic EBV syndrome, immunodeficiency); and/or 3) phenotypes that have biological relevance and analytical associations with Long COVID (depression, anxiety, diabetes, arrhythmias such as ventricular fibrillation and ventricular tachycardia,clotting disorders such as arterial thrombosis, body-mass index, lung cancer, myocardial infarction, fibromyalgia, and chronic fatigue ) based on literature review (Su et al. 2023; Thompson et al. 2023; Al-Aly, Xie, and Bowe 2021; Davis et al. 2023).

### Mendelian randomization

Based on the estimates of genetic correlation between several chronic disease phenotypes and Long COVID, we explored whether there was a potential causal role of genetic liability to these conditions and Long COVID by conducting two sample Mendelian randomization (MR) using the *twosampleMR* package (Bowden et al. 2016) in participants of European ancestry. The phenotypes that met all of the following criteria were further considered for MR analysis: 1) more than 5,000 cases in their respective GWAS analysis; 2) genetic correlations met a statistical p-value threshold of 0.002 /(0.05/26); 3) heritability of >1% as estimated by LD score regression; 4) predisposing conditions that are associated with Long COVID in prior epidemiological studies (Taylor et al. 2023; Naik et al. 2024; Ursini et al. 2021). Traits that met these four conditions were chronic fatigue, fibromyalgia, and depression. A sub-cohort of non-overlapping samples between each phenotype and Long COVID was derived and GWAS analysis was performed in participants of European ancestry only. A genetic instrument for each phenotype was derived using summary statistics. The index variants from GWAS analysis (defined as above) that had minor allele frequency of ≥0.01 were used to derive a genetic instrument for each phenotype. The estimates for MR instruments were subsequently obtained from Long COVID GWAS conducted in participants of European ancestry. We then fitted random effects inverse variance weighted (IVW) models to obtain MR estimates for each non-COVID-19 phenotype. To test the veracity of the findings, we used Steiger filtering and robust MR approaches including weighted median and MR-Egger methods (Bowden et al. 2016; Bowden, Davey Smith, and Burgess 2015; Hemani, Tilling, and Davey Smith 2017).

### Sensitivity analyses

We repeated the above GWAS analytical approach for the Long COVID Impact phenotype to identify genetic signals for a more severe form of Long COVID. Additional sensitivity analyses explored the impact of acute SARS-CoV-2 infection severity (as assessed by hospitalization) because it is a known risk factor for Long COVID. To that end, we first further adjusted the association of index variants and Long COVID from the main GWAS for COVID-19 hospitalization at the time of acute SARS-CoV2 infection. Second, using the same genetic instruments as in the main analysis, we evaluated MR effects of chronic fatigue, fibromyalgia, and depression on COVID-19 hospitalization. We further replicated our MR findings for Long COVID and COVID-19 hospitalization using summary statistics for these phenotypes from COVID-19 Host Genetics Initiative. In case of COVID-19 hospitalization, only multi-ancestry summary statistics for COVID-19 hospitalization (Cases N = 16,512, Controls N = 71321) were available via the HGI website (https://www.covid19hg.org/). Here, we first obtained the allele information, and effect estimates for variants in each of our instruments from HGI summary statistics. The variants were mapped using chromosome and position on hg38 genome build. Then we harmonized the allele information across 23andMe summary statistics of each phenotype and Long COVID HGI data. The variants that did not align for allele information were excluded from the instrument. We sought to replicate MR findings for depression from two separate external MR instruments for depression obtained using summary statistics from UK Biobank European participants (Howard et al. 2018) and from meta-analysis of three largest studies on depression that included UK BioBank, the 23andMe cohort, and the Psychiatric Genomics Consortium (Howard et al. 2019). The effect estimates for Long COVID were obtained from European GWAS conducted in this study. The depression phenotype in this UK BioBank was defined as self-reported depressive symptoms, or primary or secondary diagnosis of depressive mood disorder( ICD codes: F32—Single Episode Depression, F33—Recurrent Depression, F34—Persistent mood disorders, F38—Other mood disorders and F39—Unspecified mood disorders)(Howard et al. 2018, 2). The summary statistics and MR instruments were accessed from publicly available GWAS catalog and “MRCIEU/MRInstruments” R package respectively. We could not identify external instruments for chronic fatigue and fibromyalgia because none of the existing GWAS studies was sufficiently powered to identify genome-wide significant hits (Hajdarevic et al. 2022; Zorina-Lichtenwalter et al. 2023; Backman et al. 2021) .

## RESULTS

The selection of the participants is represented in **Figure 1**. Among 332,638 research participants who were eligible to participate in the Long COVID survey and reported acute SARS-CoV2 infection, 64,384 participants reported to have experienced Long COVID and 178,537 participants did not. After exclusions as described in **Figure 1**, we had an analytical cohort of 54,390 cases and 124,777 controls. Characteristics for unrelated cases and controls are summarized in **Table 1**, while characteristics by ancestry group are tabulated in **Supplementary Table 1**. Most participants (78.9%) in the analytical cohort were of European ancestry, followed by Latinx (16.6%) and African-American (4.5%) ancestry. Cases were more likely to be females (66.7%), non-tobacco users (37%), with a median age of 44 years [IQR=24] and mean BMI of 27.5 (SD=7.6). Compared to controls, cases were more likely to have high blood pressure (OR= 1.52, 95% CI: 1.48, 1.56), depression (OR= 1.98, 95% CI: 1.92, 2.02) , cardiometabolic conditions (OR= 1.60, 95% CI: 1.56, 1.64), and autoimmune conditions (OR= 1.55, 95% CI: 1.51, 1.59) after adjusting for age, sex, and ancestry. Among Long COVID cases, 4.7% (vs 0.005% controls) were hospitalized during acute SARS-CoV2 infection and 8.0% (vs 0.01% controls) had severe respiratory disease. When asked about the impact of Long COVID symptoms on daily life, 57.3% of Long COVID cases reported moderate to severe impact.

**Table 1:** Characteristics of 23andMe Research participants of Long COVID cohort.

|  |  |  |
| --- | --- | --- |
|  | **CASE** | **CONTROL** |
| **N** | **53,764** | **120,668** |
| **Demographics** |  |  |
| Age (median [IQR]) | 46.0[35.0,57.0] | 43.0[33.0,57.0] |
| Female N (%) | 41,324 (76.9) | 74,981 (62.1) |
| Race N (%) |  |  |
| African American | 2,234 (4.2) | 5,596 (4.6) |
| European | 42,899 (79.8) | 94,721 (78.5) |
| Latino | 8,631 (16.1) | 20,351 (16.9) |
| Education in years (mean (SD)) | 15.1 (2.4) | 15.4 (2.6) |
| BMI (median [IQR]) | 28.4 [24.3,34.0] | 27.3 [23.8,31.8] |
| Tobacco user N (%) | 21,223 (39.5) | 41,921 (35.7) |
| Two or more alcoholic drinks over past 2 weeks N (%) | 10,114 (18.8) | 28,733 (23.8) |
| **Health Conditions** |  |  |
| High blood pressure N (%) | 14,977 (27.8) | 25,507 (21.1) |
| Depression N (%) | 25,226 (46.9) | 37,578 (31.1) |
| COPD N (%) | 911 (1.7) | 840 (0.01) |
| High total cholesterol N (%) | 4,487 (8.3) | 8,054 (6.7) |
| High triglyceride N (%) | 3,207 (6.0) | 5,444 (4.5) |
| Chronic Fatigue N (%) | 3,980 (7.4) | 1,866 (1.5) |
| Fibromyalgia N (%) | 4,394 (8.2) | 2,631 (2.2) |
| Cardiometabolic condition N (%) | 23,566 (43.8) | 41,115 (34.1) |
| Autoimmune N (%) | 9,922 (18.4) | 14,053 (11.6) |
| COVID-19+ and hospitalized N (%) | 2,534 (4.7) | 617 (0.005) |
| COVID-19+ and severe respiratory disease N (%) | 4,282 (8.0) | 1,198 (0.01) |

### GWAS of Long COVID

Using approximately 110 million imputed genetic variants, we identified three loci associated with Long COVID in multi-ancestral analysis based on data from European, African-American, and Latinx ancestry groups (**Figure 2**, **Table 2**).

**Figure 2:**
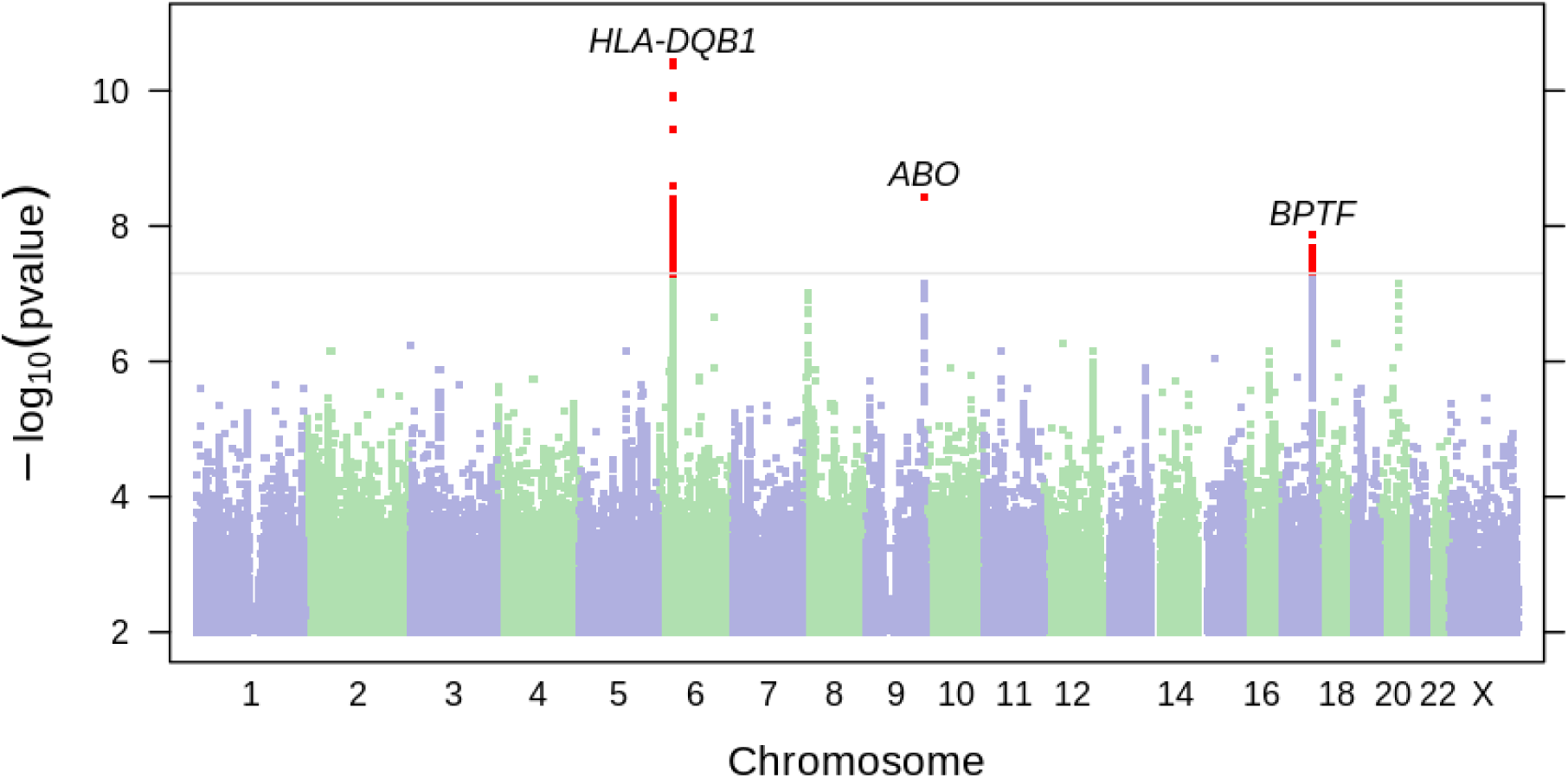
Manhattan plot of Long COVID among 23andMe participants. Manhattan plot depicts findings from the meta-analysis of three ancestral groups (European, African American, and Latinx). X-axis represents chromosomal position for each SNP. Y-axis represents negative log p-values based on logistic regression model under the additive model. Statistically significant variants are highlighted in red. The regions of associations are annotated with index variants.

**Table 2:**
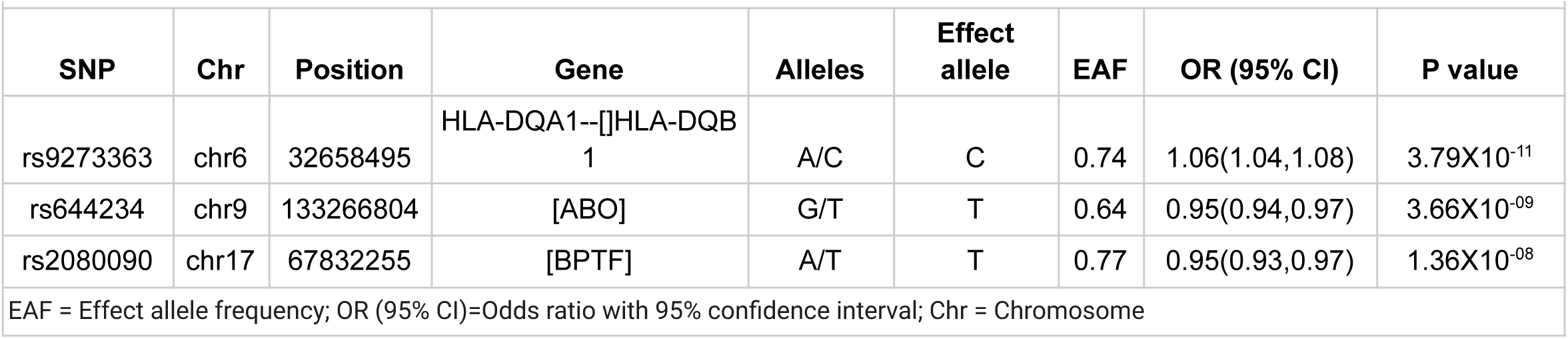
Estimates of genome-wide significant variants in meta-analysis of Long COVID GWAS.

#### Genome-wide significant loci

The top GWAS significant hit, (rs9273363; A/C with C being the effect allele, OR= 1.06, 95% CI: 1.04,1.08, p= 3.79X10^-11^) was located in chromosome 6 in the intergenic region spanning *HLA-DQA1* and *HLA-DQB1* (**Supplemental Figure 1**).The effect sizes for rs9273363 were similar across all three ancestries (p-value for heterogeneity across ancestries= 0.75). Further analysis of specific *HLA* alleles and Long COVID showed that *HLA-DRB1*\*11:04 (OR=1.18, 95% CI: 1.11, 1.24, p = 7.0 X 10^-09^), HLA-C*07:01 (OR=0.94, 95%CI = 0.92, 0.96, p = 1.14 X 10^-08^) , HLA-B*08:01 (OR=0.93, 95%CI = 0.91, 0.95, p = 1.5 X 10^-08^), and HLA-DQA1*03:01 (OR=0.95, 95% CI = 0.93, 0.97, p = 4.1 X 10^-08^) were significantly associated with Long COVID. The effect sizes were similar across three ancestries (**Supplementary Figure 2**).

The second most significant variant was observed at chromosome 9 in the ABO gene (rs644234, G/T with T being the effect allele, OR= 0.95, 95% CI: 0.94, 0.97, p= 3.66 X 10^-09^, p-value for heterogeneity across ancestries= 0.56) (**Supplementary Figure 3**).

Specifically, rs644234 is in linkage disequilibrium (LD) with a frame-shift variant (rs8176719) within the ABO gene (European r2= 0.97, African-American r2= 0.44, Latinx r2= 0.90). rs8176719 is one of the main variants determining the ABO blood group and has been previously linked to COVID-19 susceptibility and severity (Bugert et al. 2012; Severe Covid-19 GWAS Group et al. 2020; Shelton et al. 2021).

Another association was observed with rs2080090 (A/T with T being the effect allele; OR= 0.95, 95% CI: 0.93, 0.97, p= 1.36X10^-08^; p-value for heterogeneity across ancestries=0.69) located in vicinity of the *BPTF* gene on chromosome 17 (**Supplementary Figure 4**). There are at least 3 plausible causal genes in the locus; *BPTF*, *KPAN2*, and *C17orf58*. We observed eQTL signals in lymphoblast cells for a variant in LD (rs12601921, r^2^=0.98, p= 4.31X10^-89^) with rs2080090 for the *BPTF* gene (**Supplementary Table 2a**). rs2080090 was also in LD with a coding variant, rs7502307 (r^2^=0.82), located at *C17orf58* **(Supplementary Table 2b)**.

In European ancestry participants, the top associated SNP in this locus (rs78794747, A/T with T being the effect allele) was mapped to the intergenic region of *C17orf58* and *KPAN2*. rs78794747 is in high LD with multiple variants associated with gene expression in CD14 monocytes, frontal cortex, and islets of Langerhans cells (**Supplementary Table 2c).** The *C17orf58* gene has been associated with posterior myocardial infarction (Norland et al. 2019). *KPAN2* encodes nuclear transport factor importin α1, which is associated with viral suppression (Miyamoto et al. 2021). Previous studies have shown that an accessory protein encoded by the SARS-CoV2 genome,

ORF6, binds to importin α1, enhancing viral propagation by inhibiting interferon type 1 signaling (International HIV Controllers Study et al. 2010; Yuen et al. 2020; Miyamoto et al. 2021). Ancestry-specific GWAS hits are described further in **Supplemental Results** and **Supplementary Table 3a-3c**.

Associations between genome-wide significant variants and Long COVID persisted upon adjusting for COVID-19 hospitalization (**Supplementary Table 4a**). When we assessed the associations between these variants and COVID-19 hospitalization in our study adjusting for age, age*sex, and principal components of ancestry, none of them was significantly associated **(Supplementary Table 4b)**.

In the summary statistics of the HGI Long COVID GWAS including 6,450 Cases and 46,208 controls, effect sizes of our index variants were similar. However, none of the associations observed in our study was statistically significant in HGI data, which consisted of almost 10x fewer cases (**Supplementary Table 5**). When we examined the effect of rs9367106 near *FOXP4* (the significantly associated variant in HGI analyses of Long COVID cases and population controls), the association in our study was not statistically significant (p=0.57). Consistently with our observation, this variant was not significantly associated with Long COVID in the HGI data (p=0.04) when the analyses were limited to individuals who reported to be positive for SARS-CoV2.

### GWAS of Long COVID Impact

In addition to *HLA-DQA1* and *HLA-DQB1* mentioned above, we identified one indel variant on chromosome 6 in the intergenic region of *MICA* and *MICB* (OR = 0.91, 95% CI: 0.88,0.94, p = 1.18 X 10^-09^, p-heterogeneity across ancestries=0.61) (**Supplementary Figure 5, Supplementary Table 6**). The variant, rs9281499, was in LD with multiple variants that showed statistically significant pQTL signals for MICB protein in blood plasma (**Supplementary Figure 6**, **Supplementary Table 7a**). Additionally, rs9281499 is in LD with rs2596510 (r^2^=0.82) which is associated with CYP21A2 expression in type1 (p=2.09X10^-10^) and type 2 (p=2.96 X 10^-13^) T-helper cells, and CD4 T lymphocytes (2.45X10^-10^) (**Supplementary Table 7b**).

There were two novel statistically significant loci in the multi-ancestry GWAS of Long COVID Impact phenotype. The top hit, rs58970548, is an indel variant located at the intergenic region of *NMUR2* and *GRIA1* (OR= 0.94, 95% CI: 0.93, 0.96, p=4.27X10^-8^, p-heterogeneity across ancestries=0.06). (**Supplementary Figure 7**). An additional significant locus was observed at rs190759626 (G/T with T being the effect allele) located at the intergenic region of *NUTM2A* and *NUTM2D* (**Supplementary Figure 8**) on chromosome 10. Ancestry specific analyses are reported in **Supplementary Results**.

### Genetic Correlation with Similar Phenotypes

We identified 26 conditions in participants of European ancestry that met the criteria of shared pathophysiology, similar clinical features, and previously published associations with Long COVID **Supplementary Table 8** (Sherif et al. 2023; Davis et al. 2023; Peter et al. 2022; Cohen et al. 2022; Bull-Otterson et al. 2022; Davis et al. 2021; Ursini et al. 2021). We further limited the list of conditions to those with at least 5,000 cases contributing to GWAS, p-value < 0.002 (0.05/26) for the genetic correlation with Long COVID, and estimated heritability more than 1%. Overall, 13 phenotypes met the criteria and are presented in **Figure 3**. The strongest genetic correlations were observed between Long COVID and chronic pain (r_g_=0.72,p=2.49X10^-170^), chronic fatigue (r_g_=0.71,p=1.09X10^-120^), and fibromyalgia (r_g_=0.70,p=6.30X10^-142^).

**Figure 3:**
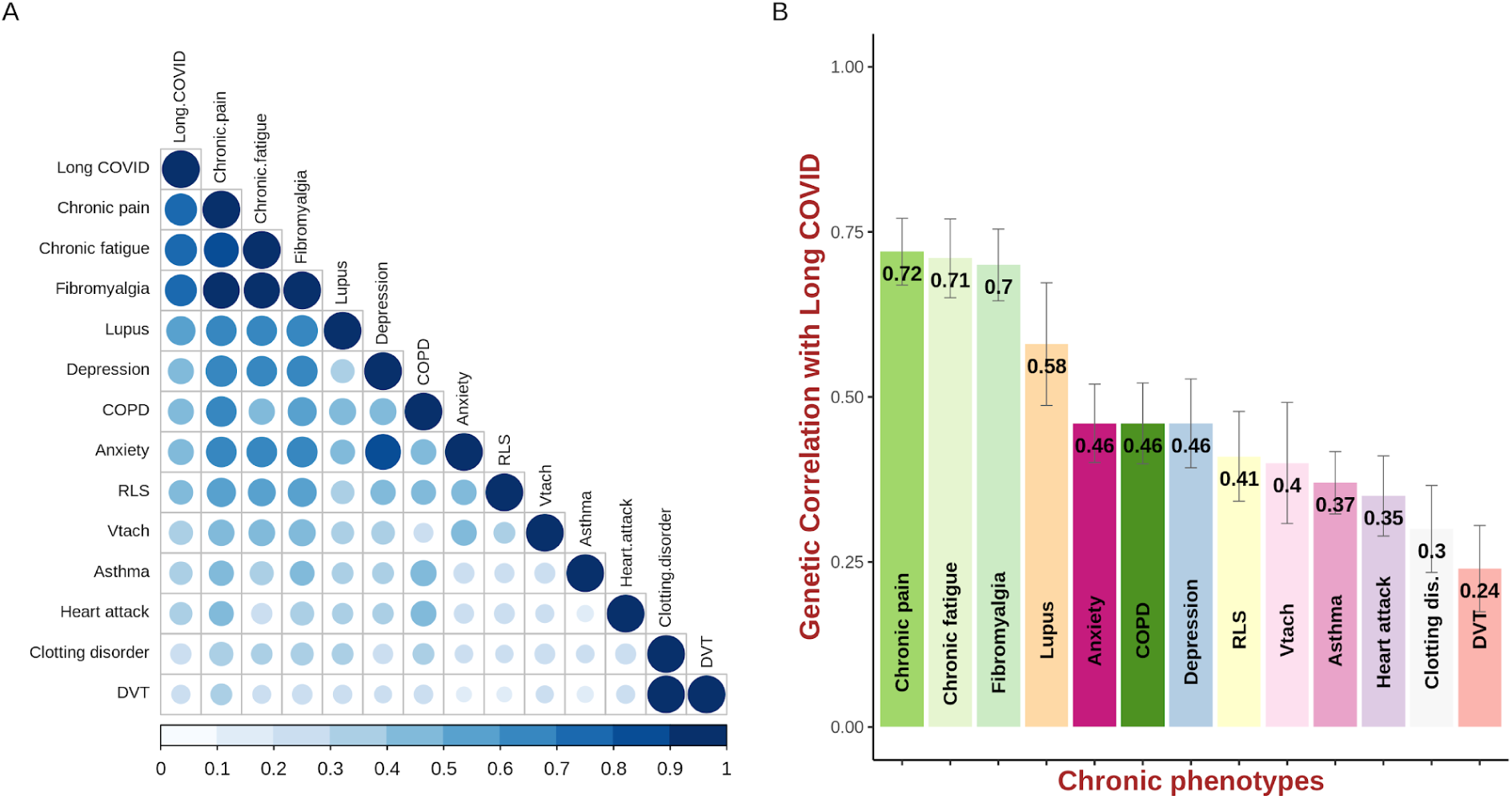
Genetic correlation of Long COVID with chronic phenotypes in 23andMe participants of European ancestry. **Panel A:** Heat map showing genetic correlations between chronic phenotypes, including Long COVID, obtained from LD Score regression. **Panel B**: Bar plot represents genetic correlation values of chronic phenotypes and Long COVID obtained from LD Score regression. X-axis shows a list of chronic phenotypes plotted. Y-axis shows values of genetic correlation from 0 to 1. A complete list of phenotypes are included in Supplementary Table 8. RLS = Restless leg syndrome; Vtach = Ventricular tachycardia; DVT = Deep vein thrombosis; dis. = disorders; COPD = Chronic obstructive pulmonary disease

### Mendelian Randomization

We investigated the potential causal role of three phenotypes (chronic fatigue, fibromyalgia, and depression) with Long COVID through Mendelian randomization (MR). We did not consider chronic pain due to its nonspecific definition in the 23andMe database. The genetic instrument for chronic fatigue included 186 SNPs (mean F-statistic per SNP = 43.17), for fibromyalgia included 349 SNPs (mean F-statistic per SNP=45.04), and for depression included 696 SNPs (mean F-statistic per SNP=53.8). We found strong evidence of a potential causal effect to each of these three conditions on Long COVID (chronic fatigue: OR=1.59 (95%CI: 1.51, 1.66), fibromyalgia: OR=1.54 (95%CI: 1.49, 1.60), and depression: OR=1.53 (95%CI: 1.46, 1.61); estimates from IVW-MR). These effects persisted when employing robust MR approaches including weighted median and MR Egger (**Figure 4, Supplementary Figure 9, Supplementary Table 9a**). Steiger filtered and outlier filtered estimates (**Supplementary Table 9a**) were similar to the respective IVW estimates for chronic fatigue and fibromyalgia. However, for depression, the Steiger filtered estimates were closer to the null than IVW estimates. The MR-Egger intercept for fibromyalgia was non-zero (-0.01, p = 0.01) with the resultant MR-Egger causal estimate being stronger than IVW (**Supplementary Table 9a**). The Mendelian randomization findings were replicated and were in a direction similar to our findings when the genetic instruments for the exposures derived from the 23andMe research cohort were applied to disease estimates derived from the Long COVID HGI consortium (**Supplementary Figure 10, Supplementary Table 10a**). In the Mendelian randomization analyses, the directions of effects for exposures applied to COVID-19 hospitalization data from the COVID HGI consortium were also similar to our findings (**Supplementary Table 10b**, **Supplementary Figure 11**). When we repeated Mendelian randomization using the Long COVID Impact definition, the results were consistent (**Figure 4, Supplementary Figure 12, Supplementary Table 9b**). MR analysis showed evidence of genetic liability to chronic fatigue syndrome, fibromyalgia, and depression leading to a higher risk of COVID-19 hospitalization (**Figure 4, Supplementary Figure 13, Supplementary Table 9c**). Our results were also consistent after using external instruments for depression derived solely from UK Biobank GWAS which included 35 SNPs with a mean F statistic of 40.83, and instruments derived from a meta-analysis of the UK BioBank, the 23andMe cohort, and the Psychiatric Genomics Consortium including 87 SNPs with mean F statistic of 77.67 (**Supplementary Figure 14**).

**Figure 4:**
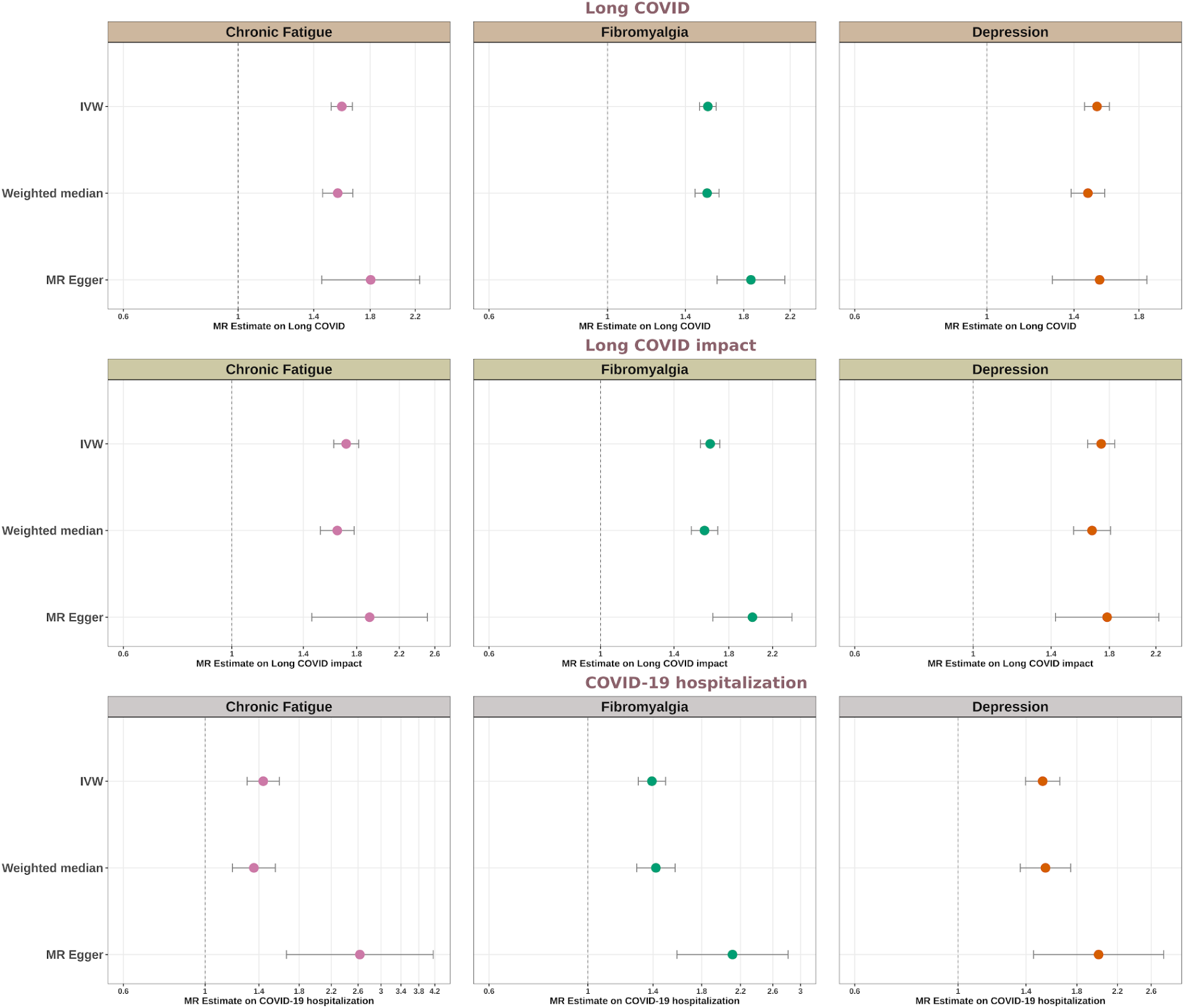
Forest plot representing genetically predicted effects of chronic phenotypes using Mendelian randomization. Top row panel: *Genetically predicted effects of chronic phenotypes on Long COVID* Middle row panel: *Genetically predicted effects of chronic phenotypes on Long COVID impact* Bottom row panel: *Genetically predicted effects of chronic phenotypes on COVID-19 hospitalization* The estimates in the plot depict odds ratio and 95% confidence intervals. The genetic instrument for chronic fatigue is derived using information from 186 SNPs (mean F statistic = 43.17). The genetic instrument for chronic fatigue is derived using information from 349 SNPs (mean F statistic = 45.04). The genetic instrument for depression is derived using information from 696 SNPs (mean F statistic = 53.8). IVW = Inverse variance weighted.

## DISCUSSION

We report the largest GWAS of Long COVID to date among ancestrally diverse participants with a history of SARS-CoV2 acute infection. Our top finding links the *HLA* region, in particular the *HLA-DRB1*\*11:04 variant, to developing Long COVID. Additional hits were observed in *ABO*, a previously reported severity locus, and a novel region spanning the *BPTF-C17orf58-KPNA2* genes. Genetic correlation analysis showed strong relationships between Long COVID and multiple chronic conditions, including chronic fatigue, depression, and fibromyalgia. We identified strong evidence of a potential causal effect of genetic liability to these conditions on the risk of Long COVID.

To date, only one study has reported GWAS findings of Long COVID, conducted by the COVID-19 Host Genetics Initiative (HGI) (Lammi et al. 2023). The HGI study compared 6,450 Long COVID cases with 1,093,995 population controls from multiple countries and identified a genome-wide significant association with rs9367106 in the *FOXP4* locus. In contrast, our study has approximately a 7-fold larger number of cases, and is contained within one cohort, increasing statistical power through sample size and removing between-study heterogeneity, respectively. The association detected in the COVID-19 HGI study was not significant in our analysis; cases and controls in our study were both infected with SARS-CoV2, removing the effects of COVID-19 susceptibility by design. The *FOXP4* locus identified by HGI was functionally mapped to COVID-19 severity and lung function (Lammi et al. 2023; Wu et al. 2021; Pairo-Castineira et al. 2023), and potentially reflects the genetic signal relevant to acute infection susceptibility and/or severity. In contrast, our analysis identified multiple loci (*HLA-DRB1*–*HLA-DQA1*, *BPTF*) that are independent of SARS-CoV-2 hospitalization, another marker of COVID-19 severity, and instead are enriched for immune pathways. Assessing the genome-wide significant hits of our study in the HGI data, the direction and effect sizes for these variants were similar but did not meet statistical significance, likely because our study was powered to pick up more subtle signals (**Supplemental Table 5**).

We observed associations of Long COVID with variants in the *HLA* locus, highlighting the role of immune mechanisms as postulated in earlier studies (Klein et al. 2023; Davis et al. 2023; Augusto et al. 2023). *HLA* class II molecules including our top hit, *HLA-DRB1*, play a critical role in the adaptive immune system by presenting foreign peptide antigens to receptors on T lymphocytes, initiating an immune response on exposure to viral antigens. Specifically, *HLA-DRB1*\*11 alleles have been previously associated with asymptomatic SARS-CoV2 infections (Augusto et al. 2023; Ouedraogo et al. 2023). While the role of *HLA* variation in Long COVID has not been extensively studied, a prior smaller immunoprofiling study has found monocyte expression of MHC class II (HLA-DR) to be higher among Long COVID cases compared to controls who had experienced an acute SARS-CoV2 expression and recovered (Klein et al. 2023). The discovery of the *HLA* association in a hypothesis-free analysis underscores the importance of antigen presentation in the genetic susceptibility to Long COVID, potentially informing future risk stratification and treatment approaches.

Our study contributes to the body of evidence in support of the role of *ABO* loci in SARS-CoV2 infection outcomes. Previous studies have identified *ABO* loci as independently associated with COVID-19 susceptibility and severity (Shelton et al. 2021; Severe Covid-19 GWAS Group et al. 2020; Pairo-Castineira et al. 2023; Zhao et al. 2021). While differential susceptibility to SARS-CoV2 infection was controlled for in the design of our study, acute infection severity is a well-established risk factor for long COVID (Tsampasian et al. 2023). The association between *ABO* variation and COVID-19 outcomes is potentially driven by the ABO antibodies interfering with the attachment of viral spike proteins to ACE2 receptors (Breiman, Ruvën-Clouet, and Le Pendu 2020; Guillon et al. 2008). Additionally, a proteomic analysis of rs657152 (LD r^2^ = 0.99 with our GWAS hit rs644234) within the *ABO* locus in a population-based cohort identified proteins related to immune function such as IL-15 and STAT3 signaling (Steffen et al. 2022). IL-15 has been proposed as a therapeutic strategy to improve acute COVID-19 outcomes (Lu et al. 2022; National Library of Medicine (U.S.) 2020; Kandikattu et al. 2020). Blood group subtypes encoded by *ABO* are also associated with differential risk of thrombosis (Vasan et al. 2016), which represents another potentially relevant etiologic mechanism linking *ABO* and Long COVID. In recent studies, thrombo-inflammation has been reported as a clinical feature of Long COVID, likely driven by dysregulated coagulation mechanisms in response to viral antigen-antibody complexes (Cervia-Hasler et al. 2024).

We provide novel evidence establishing potential causal relationships between Long COVID and genetically correlated conditions that share similar symptomologies, namely chronic fatigue, depression, and fibromyalgia. The findings that genetic liability to chronic fatigue, depression and fibromyalgia also leads to a higher risk of hospitalization for acute SARS-CoV-2 infection provides mechanistic insights. Prior studies have reported depression can impair function of a broad range of immune cells (Drevets et al. 2022; Walther et al. 2022). While the relationship of chronic fatigue and fibromyalgia with immune dysfunction is less clear, NK cell activity may be impaired among these individuals (Brenu et al. 2012; Rivas et al. 2018; Verma et al. 2022), with altered NK cell function also identified among patients with acute severe COVID-19 (Malengier-Devlies et al. 2022). Furthermore, there is growing evidence that immune and oxidative stress pathways induce neuroaffective effects resulting in symptoms such as chronic fatigue syndrome and depression (Al-Hakeim et al. 2023). A recent study found that Long COVID and chronic fatigue syndrome share a common picture of impaired immune response secondary to metabolic consequences of elevated oxidative stress in T lymphocytes (Shankar et al. 2024). Thus the immune impact of chronic fatigue, depression, and fibromyalgia potentially explains the well-documented higher risk of Long COVID among individuals who experienced severe acute COVID-19. While the immune pathway likely doesn’t fully account for the increased risk of Long COVID, it provides new information that could help identify subgroups of individuals that ought to receive preventative measures such as ongoing vaccination to avoid severe acute infection. Also notable from a translational perspective is the indel variant, rs58970548, near the *GRIA1* gene in the GWAS of the Long COVID impact phenotype. Though we could not identify any functional signals at predetermined thresholds of significance (r^2^>0.80 and p < 5 X 10^-08^) in our datasets, the variants within 500kb and in linkage disequilibrium (r^2^>0.5) of rs58970548 have been associated with various psychiatric conditions (Parekh et al. 2018; Ismail et al. 2022; Chiesa et al. 2012). Specifically, *GRIA1* has been recently identified in brain proteome data as a potential drug target for depression (Liu et al. 2023).

Our study has multiple strengths and limitations. First, this is the largest GWAS study of Long COVID to date that identified multiple etiologically relevant loci that help shed light on Long COVID biology. Given the constellation of symptoms of Long COVID and their varying severity at individual level (Davis et al. 2021), setting clinical trials for Long COVID has been challenging in the absence of such data on biological mechanisms. By providing evidence of immune function being a likely driving force for etiological tapestry of Long COVID and associated conditions, there is an opportunity to reassess the feasibility of repurposing existing therapeutic interventions planned in the Long COVID trials. Second, the extensive database of 23andMe provided a unique opportunity to study the shared genetic architecture of multiple phenotypes, linking Long COVID with other disorders characterized by substantial unmet clinical need such as chronic fatigue syndrome, depression, and fibromyalgia. Third, owing to the inclusion of more genome-wide significant SNPs compared to prior studies (Lammi et al. 2023), the genetic instruments explain a greater proportion of the genetic variability in the exposures, potentially leading to more reliable MR estimates. Our MR estimates were successfully replicated in an external dataset, lending further credence to our causal conclusions. However, our findings must be interpreted in the context of several limitations. The use of self-reported data enabled us to capture cases that may have been overlooked due to diagnostic challenges surrounding Long COVID (Knuppel et al. 2023). Nevertheless, the self-reported data may also have introduced misclassification bias due to subjectivity in grading of symptoms at the individual level. Additionally, because participation in 23andMe surveys is self-selected, our findings may not be representative of the broader population. Furthermore, collider bias is a recognized threat to the validity of COVID-19 studies (Griffith et al. 2020). However, the 696 genetic variants comprising the MR instrument for depression were not associated with participation into our study (p= 0.82) and almost all study participants responding to a survey reported having experienced acute COVID-19, making both study respondents to the Long Covid survey and study participation on the basis of previous acute COVID-19 infection unlikely colliders, collectively alleviating selection bias concerns.

In summary, the largest multi-ancestry GWAS of Long COVID to date highlights new biological mechanisms independent of COVID-19 susceptibility. Furthermore, we report robust genetic correlations between Long COVID and a number of phenotypes with similar symptoms, including chronic fatigue, fibromyalgia, and depression, as well as a causal effect of liability to these conditions on Long COVID. Together, these findings can help identify at-risk individuals for Long COVID, as well as provide novel insights that support developing therapeutic options for both Long COVID and symptomatically similar conditions.

## Supporting information

Supplementary Results

## ACKNOWLEDGEMENTS

We would like to thank the research participants and employees of 23andMe for making this work possible. The following members of the 23andMe Research Team contributed to this study:

Stella Aslibekyan, Adam Auton, Elizabeth Babalola, Robert K. Bell, Jessica Bielenberg, Ninad S. Chaudhary, Zayn Cochinwala, Sayantan Das, Emily DelloRusso, Payam Dibaeinia, Sarah L. Elson, Nicholas Eriksson, Chris Eijsbouts, Teresa Filshtein, Pierre Fontanillas, Davide Foletti, Will Freyman, Zach Fuller, Julie M. Granka, Chris German, Éadaoin Harney, Alejandro Hernandez, Barry Hicks, David A. Hinds, M. Reza Jabalameli, Ethan M. Jewett, Yunxuan Jiang, Sotiris Karagounis, Lucy Kaufmann, Matt Kmiecik, Katelyn Kukar, Alan Kwong, Keng-Han Lin, Yanyu Liang, Bianca A. Llamas, Aly Khan, Steven J. Micheletti, Matthew H. McIntyre, Meghan E. Moreno, Priyanka Nandakumar, Dominique T. Nguyen, Jared O’Connell, Steve Pitts, G. David Poznik, Alexandra Reynoso, Shubham Saini, Morgan Schumacher, Leah Selcer, Anjali J. Shastri, Jingchunzi Shi, Suyash Shringarpure, Keaton Stagaman, Teague Sterling, Qiaojuan Jane Su, Joyce Y. Tung, Susana A. Tat, Vinh Tran, Xin Wang, Wei Wang, Catherine H. Weldon, Amy L. Williams, Peter Wilton.

## Author Contributions

The 23andMe COVID-19 Team developed the recruitment and participant engagement strategy and acquired and processed the data. N.S.C analyzed the data. N.S.C., P.A, C.H.W, S.A, and M.V.H interpreted the data. N.S.C., S.A. and M.V.H. wrote the manuscript. All authors participated in the preparation of the manuscript by reading and commenting on the drafts before submission.

