## Supplementary Results for "Multi-ancestry GWAS of Long COVID identifies immune-related loci and etiological links to chronic fatigue syndrome, fibromyalgia and depression"

***Ancestry specific loci in the GWAS of Long COVID phenotype***

The top significant variant in GWAS of participants of European ancestry was rs41291806 (A/C with C being the effect allele, p = 2.9 X $10^{-10}$) located in the intergenic region of *HLA-DRA* and *HLA-DRB5* (**Supplementary Table 3a**). The remaining two loci from the GWAS of participants of European ancestry were rs898642871 (A/G with G being the effect allele, p = 4.51 X $10^{-09}$ ) near *HNF4G* gene on chromosome 8 and rs919530877 (C/T with T being the effect allele, p = 6.39 X $10^{-09}$) located in the intergenic region of *POU3F1* and *RRAGC.* Both these variants are ultra rare (MAF <0.0001) in the European population. The genes, *POU3F1* and *RRAGC,* are primarily involved in positive regulation of RNA Polymerase II and MTOR signaling. However, these two low-frequency variants are the only variants meeting the threshold in the respective regions of significance and can be false positive associations. The variant, rs192518628 (C/T with T being the effect allele, p = 2.9 X $10^{-08}$, minor allele frequency= 0.017) near *LRRC4C* gene has no known biological role. Only one variant was genome-wide significant in participants of African ancestry (**Supplementary Table 3b**). rs78314263 (A/G with G being the effect allele, p = 3.4X$10^{-08}$) is an intronic variant located on *CALN1* gene which encodes for calcium-binding proteins and is associated with the nervous system. Among latinx participants, the variant (rs117011610, C/G with G being the effect allele, p = 4.16 X $10^{-08}$) on chromosome 15 at *MYO9A* gene met the statistical threshold (**Supplementary Table 3c**).

***Ancestry specific loci in the GWAS of Long COVID Impact phenotype***

Compared to trans-ancestry analysis, there were significant functional associations on ancestry specific GWAS of Long COVID Impact of European participants (**Supplementary Table 7d**). The most significant variant, rs9265958 (A/G with G being the effect allele, p=3.15X$10^{-10}$ ), is located at the *HLA* region between *HLA-C* and *HLA-B* and is in LD with three missense variants: rs7750641 (r2=0.86, gene=*TCF19*), rs2596492 (r2=0.84, gene=*HLA-B*), and rs3134900 (r2=0.81,gene=*MICB*). Additionally, rs9267488 (r2=0.85) is a splice variant on gene *ATP6V1G2* which is associated with ATP-ase dependent acidification of intracellular components that plays a part in protein sorting and receptor-mediated endocytosis [1]. Most of the significant expressions on eQTL analysis for rs9265958 were associated with naive CD4+ T cells (eQTL genes: *FLOT1*, *IER3*, *VARS2, CYP21A2*) , naive CD8+ T cell (eQTL gene: *CLIC-1*), and T-helper cells (eQTL genes: *C4B, CYP21A2, HLA-B*). Additional eQTL and pQTL results are provided in **Supplementary Table 7e and 7f**). We also observed *HLA* allelic associations with rs9265958 at *HLA-DRB1**11:04 (OR[95%CI] = 1.27 [1.18 ,1.36] , p = 2.6X$10^{-10}$) and *HLA-B**08:01 (OR[95%CI] = 0.91[0.88,0.94],p = 6.0X$10^{-09}$). Additional seven genome-wide significant loci were observed in GWAS of participants of European ancestry of which only one variant (rs190759626) had minor allele frequency more than 0.02 (**Supplementary Table 11a**). Similar to Long COVID GWAS in the European population, low-frequency of these variants was likely driving the genome-wide significance. The significant variant, rs190759626, on chromosome 10 (G/T with T being the effect allele, p = 4.0 X $10^{-09}$) also reached genome-wide significance on trans-ancestry analysis (p-heterogeneity=0.21). Only one variant was genome-wide significant among participants of African ancestry. The variant, rs192015048 (C/T with T being the effect allele, $p = 2.16 \times 10^{-08}$) , on chromosome 14 is located near the *RPS29* gene which is associated with Diamond-Blackfan anemia. (**Supplementary Table 11b**). No significant effects were noted for the variant in the functional analysis. None of the variants were significant in GWAS of participants of Latinx ancestry.

Two genome-wide significant variants for Long COVID impact GWAS were present in Long COVID genetic consortium summary statistics. These variants had similar direction and effect sizes but were not statistically significant in consortium data (**Supplementary Table 12)**.
