## Supplementary Figures for "Multi-ancestry GWAS of Long COVID identifies immune-related loci and etiological links to chronic fatigue syndrome, fibromyalgia and depression"

**Supplementary Figure 1: Regional plot around *HLA-DQA1–HLA-DQB1* locus for Long COVID**

The colors indicate the strength of LD relative to the index variant (rs9273363). The index variant is represented by gray color. Imputed variants are indicated with '+' symbols or '×' symbols for coding variants. Directly genotyped variants are indicated by 'o' symbols or diamond symbols for coding variants.

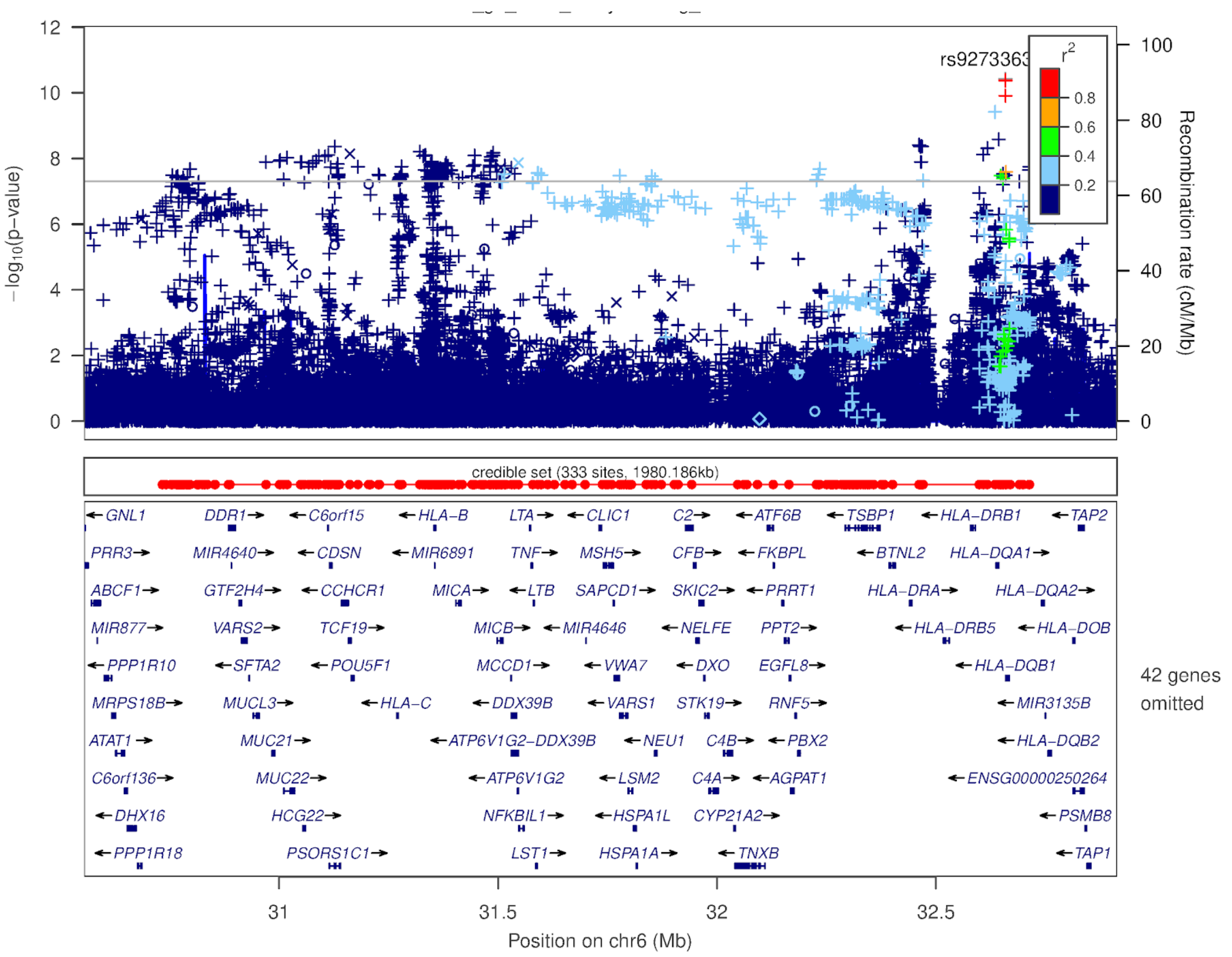

**Supplementary Figure 2: Forest plots representing meta-analysis estimates for four HLA alleles associated with Long COVID**

The fixed effect model estimates in the plot depict odds ratios and 95% confidence intervals for each ancestry (European, Latino, and African-American) and multi-ancestry meta-analysis. The meta-analysis estimates are represented by diamond. The ancestry specific estimates are presented by square. The size of the square represents the contribution of a particular ancestry towards meta-analysis.

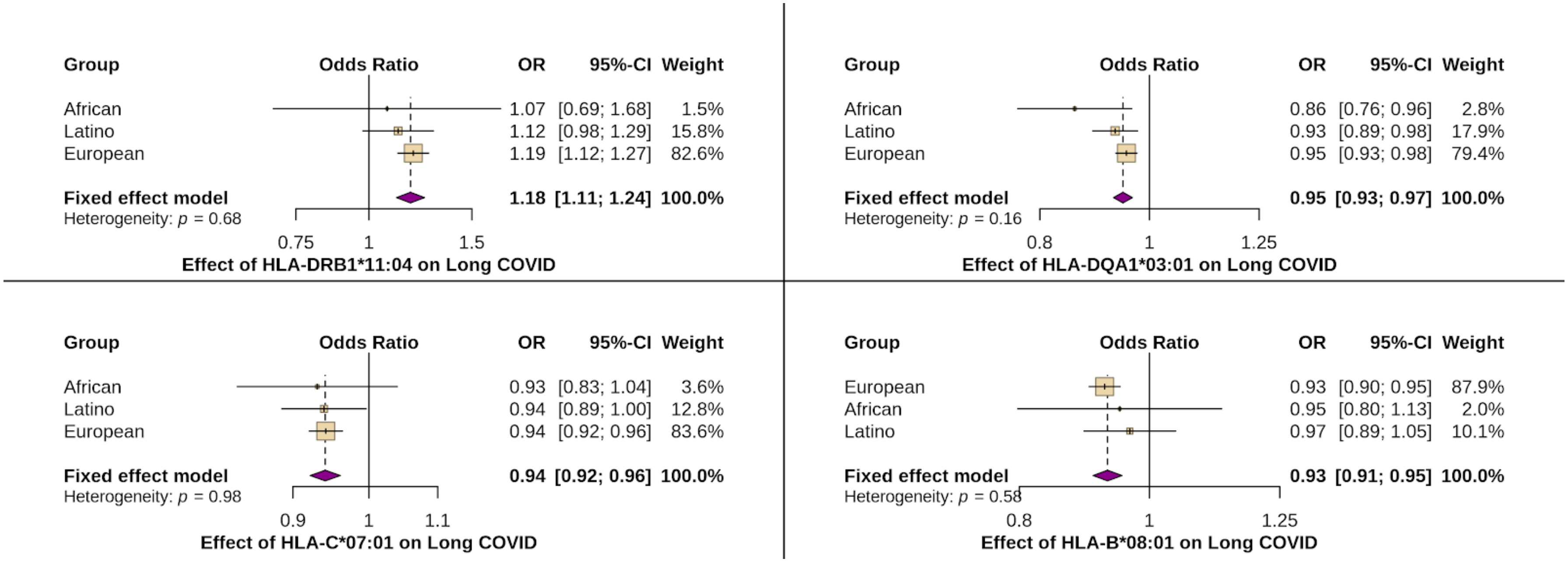

**Supplementary Figure 3: Regional plot around *ABO* locus for Long COVID**

The colors indicate the strength of LD relative to the index variant (rs644234). The index variant is represented by gray color. Imputed variants are indicated with '+' symbols or '×' symbols for coding variants. Directly genotyped variants are indicated by 'o' symbols or diamond symbols for coding variants.

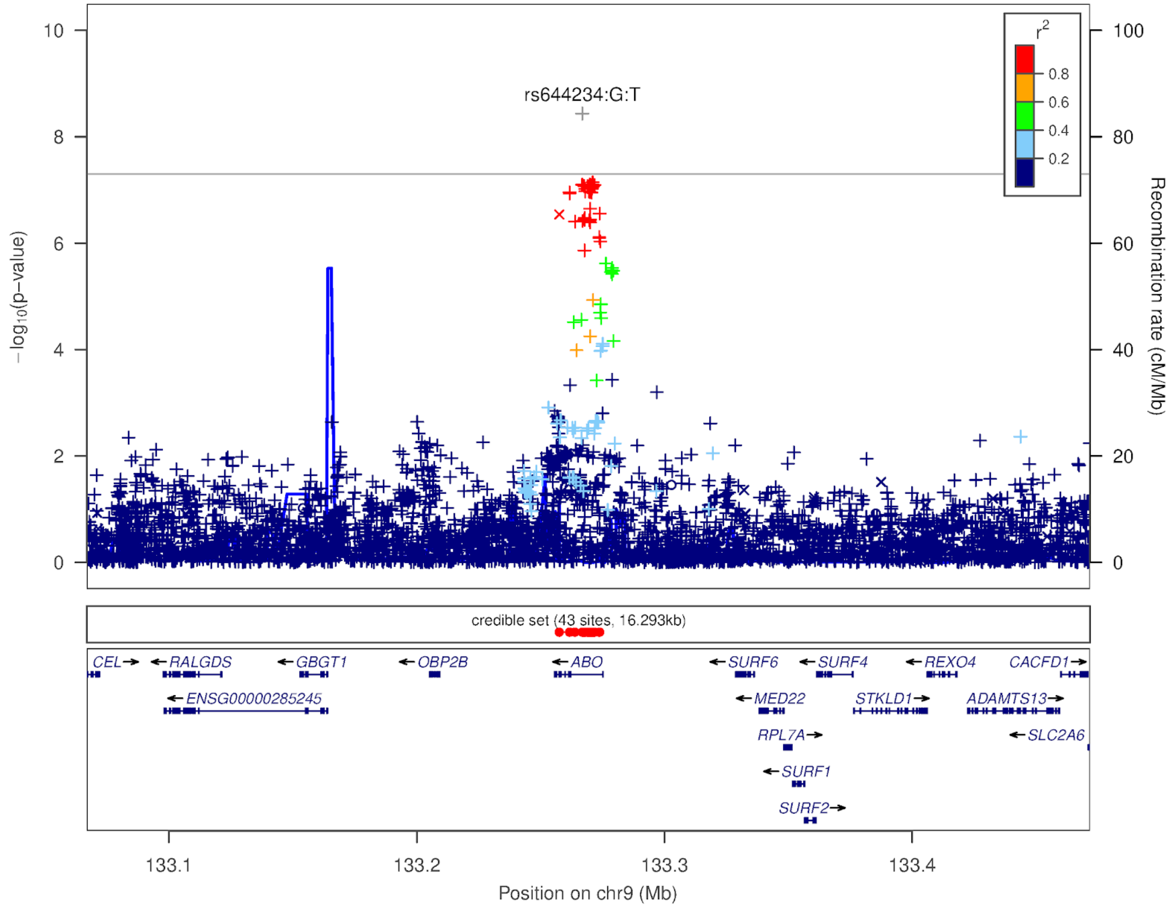

**Supplementary Figure 4: Regional plot around *BPTF* locus for Long COVID**

The colors indicate the strength of LD relative to the index variant (rs2080090). The index variant is represented by gray color. Imputed variants are indicated with '+' symbols or '×' symbols for coding variants. Directly genotyped variants are indicated by 'o' symbols or diamond symbols for coding variants.

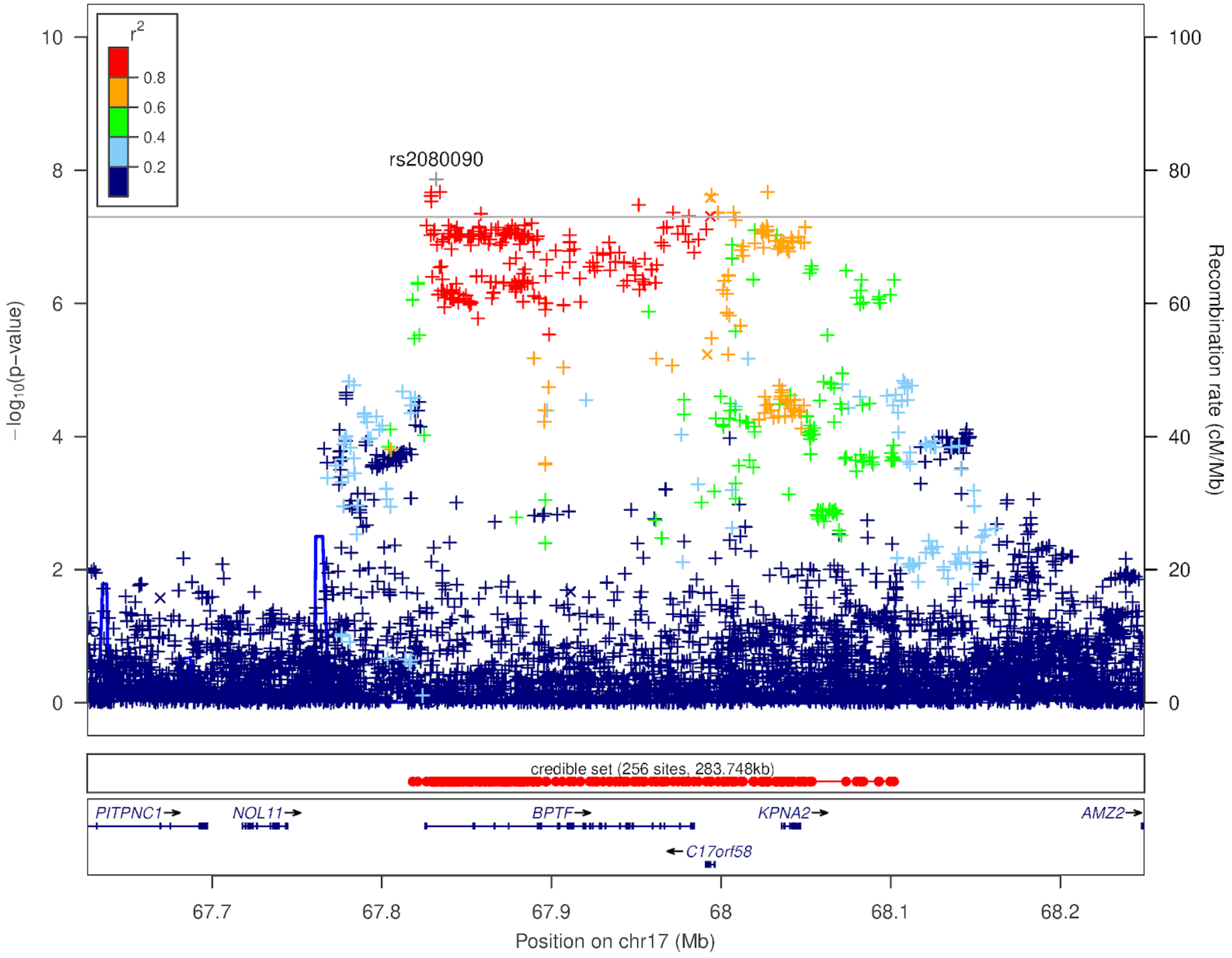

**Supplementary Figure 5: Manhattan plot of Long COVID Impact phenotype among 23andMe participants**
Manhattan plot depicts findings from the meta-analysis of three ancestral groups (European, African American, and Latinx). X-axis represents chromosomal position for each SNP. Y-axis represents negative log p-values based on logistic regression model under the additive model. Statistically significant variants are highlighted in red. The regions of associations are annotated with index variants.

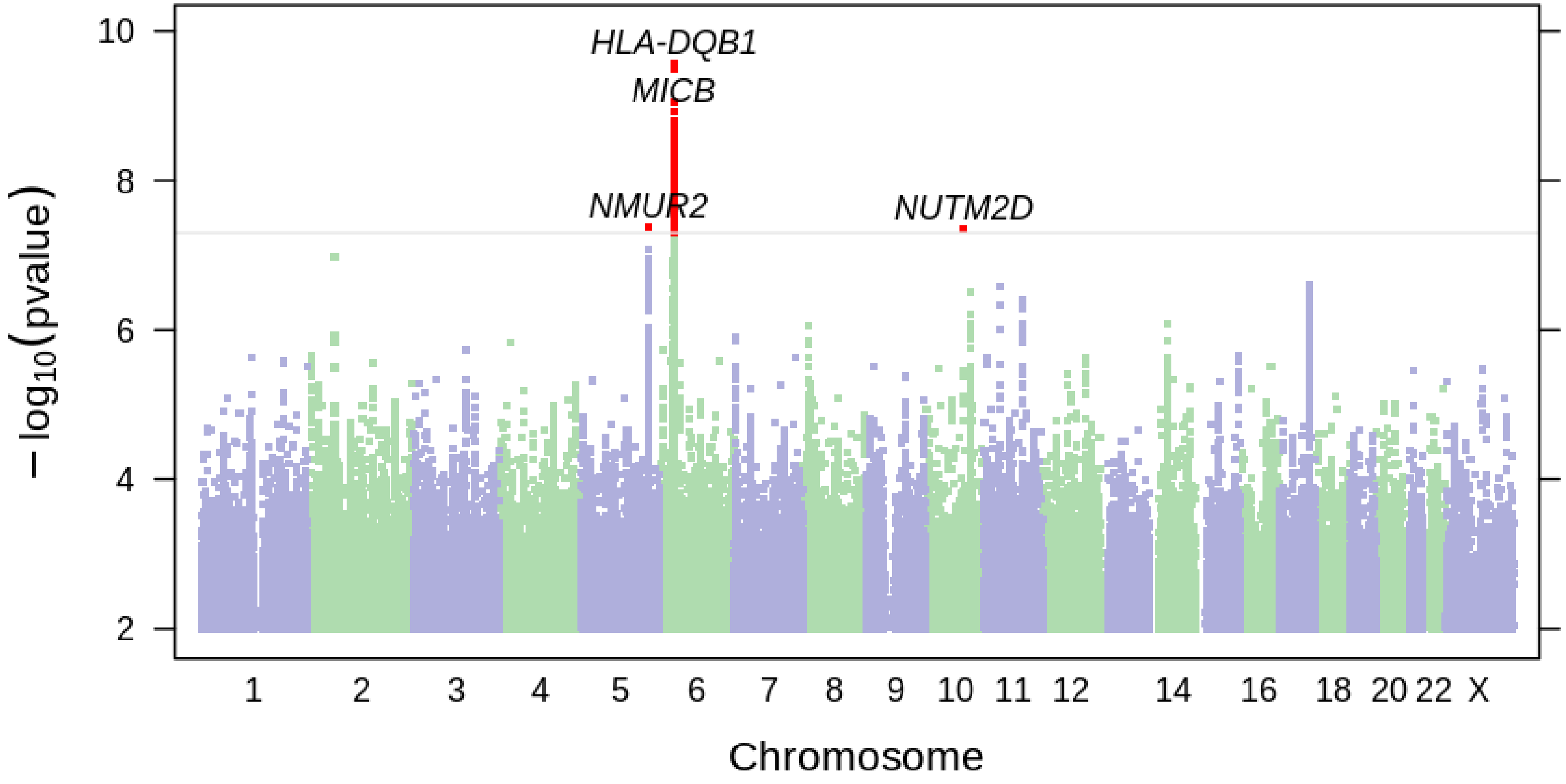

**Supplementary Figure 6: Regional plot around *MICB* locus for Long COVID Impact**

The colors indicate the strength of LD relative to the index variant (rs9281499). The index variant is represented by gray color. Imputed variants are indicated with '+' symbols or '×' symbols for coding variants. Directly genotyped variants are indicated by 'o' symbols or diamond symbols for coding variants.

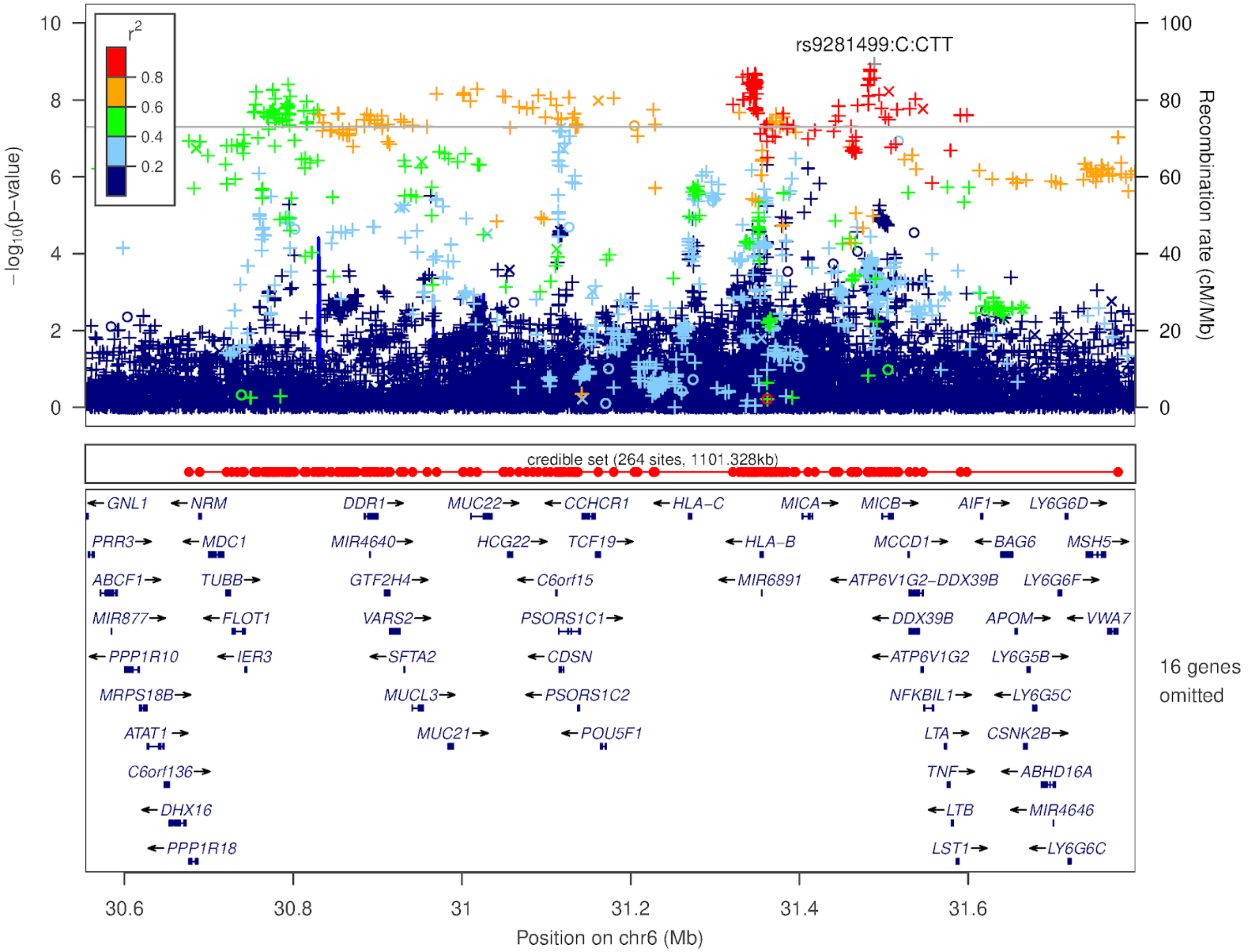

**Supplementary Figure 7: Regional plot around *NMUR2 and GRIA1* locus for Long COVID Impact**

The colors indicate the strength of LD relative to the index variant (rs58970548). The index variant is represented by gray color. Imputed variants are indicated with '+' symbols or '×' symbols for coding variants. Directly genotyped variants are indicated by 'o' symbols or diamond symbols for coding variants.

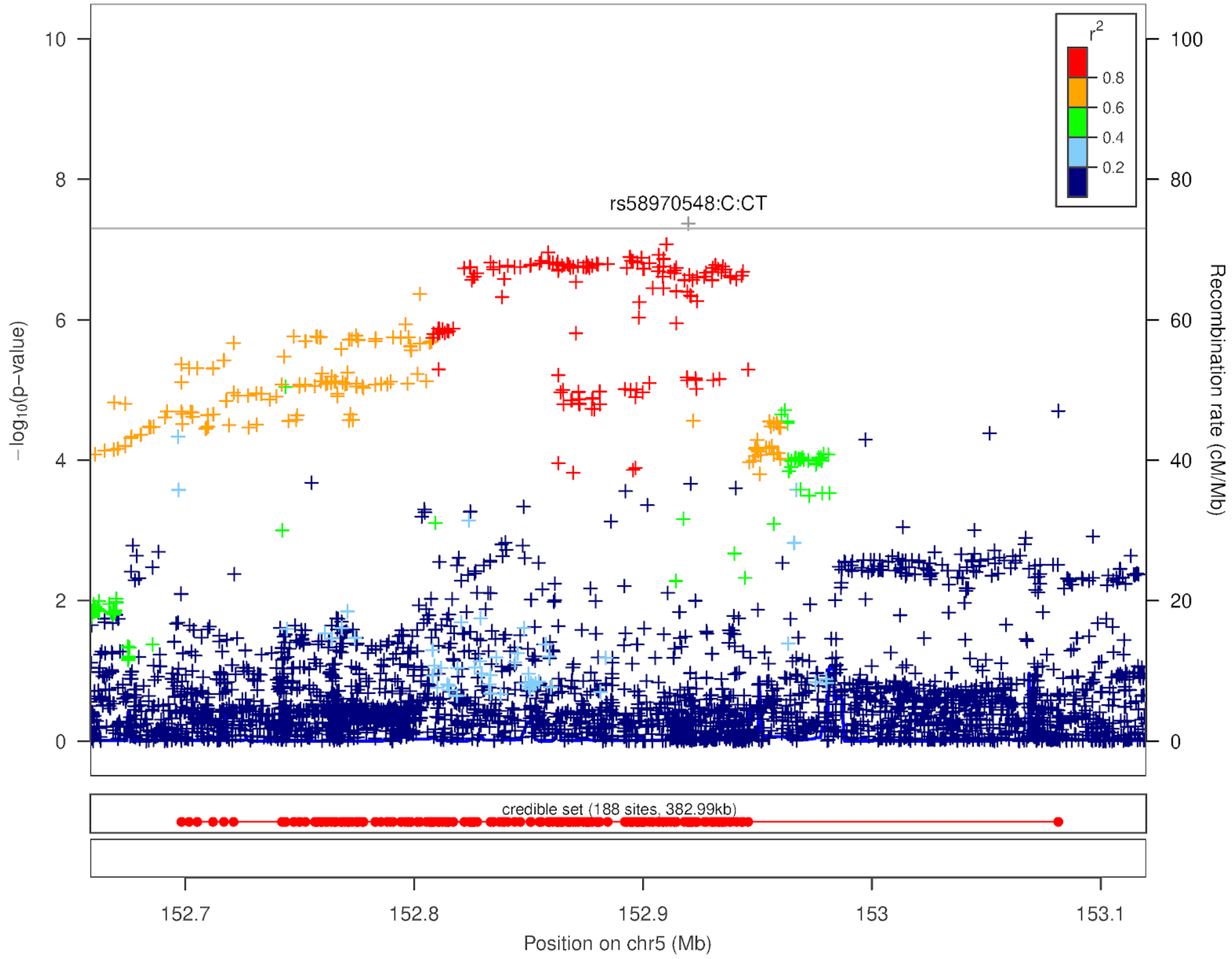

**Supplementary Figure 8: Regional plot around *NUTM2A and NUTM2D* locus for Long COVID**

The colors indicate the strength of LD relative to the index variant (rs190759626). The index variant is represented by gray color. Imputed variants are indicated with '+' symbols or '×' symbols for coding variants. Directly genotyped variants are indicated by 'o' symbols or diamond symbols for coding variants.

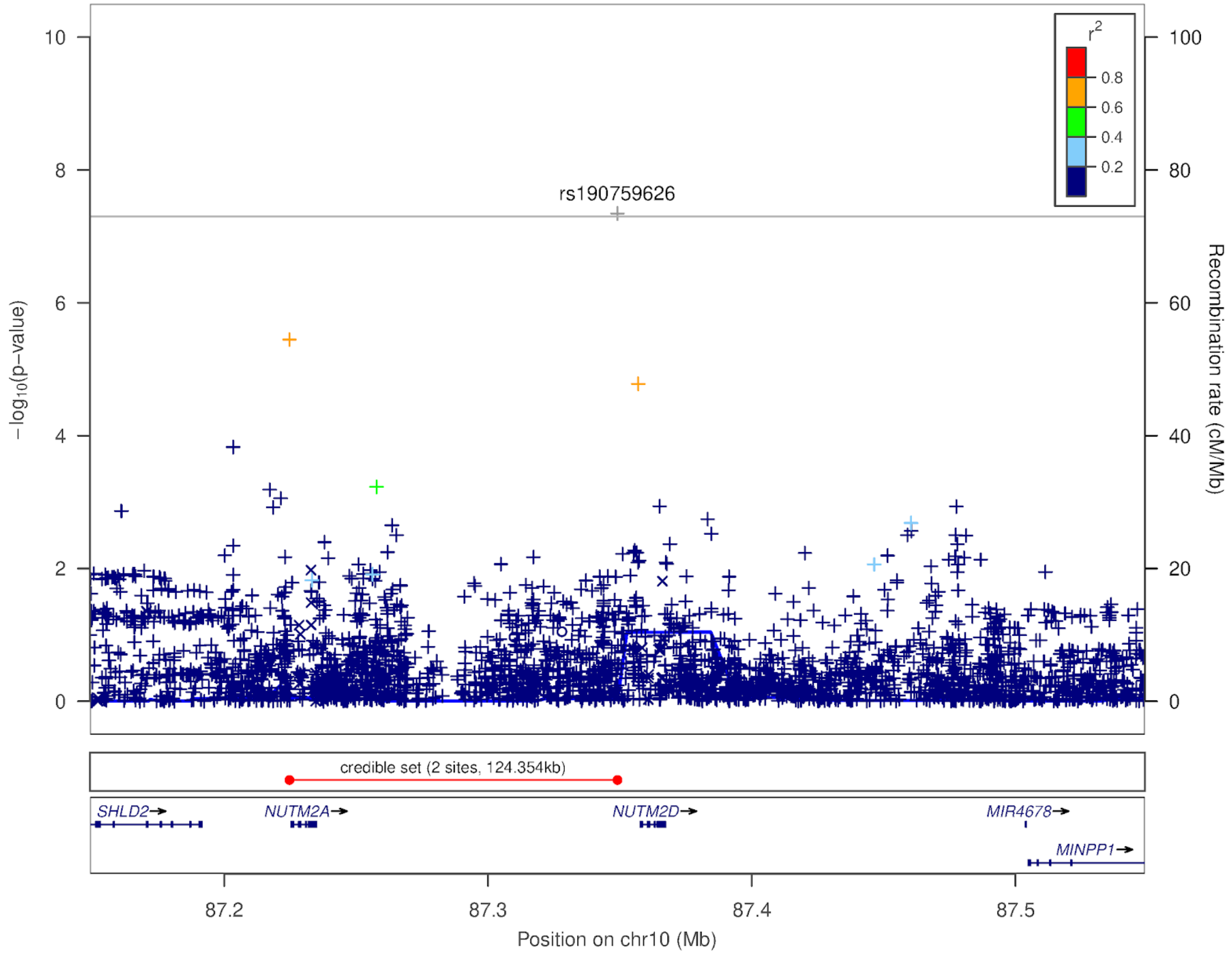

**Supplementary Figure 9: MR scatterplot of genetic associations between chronic phenotypes and Long COVID**
Panel A**:** The genetic association and corresponding 95% confidence interval (CI) for each SNP (n = 186) with chronic fatigue (x-axis) and Long COVID (y-axis) are plotted. The plot represents estimates from three methods: random-effects inverse weighted variance method, weighted median, and random-effects MR Egger.
Panel B**:** The genetic association and corresponding 95% confidence interval (CI) for each SNP (n = 349) with fibromyalgia (x-axis) and Long COVID (y-axis) are plotted. The plot represents estimates from three methods: random-effects inverse weighted variance method, weighted median, and random-effects MR Egger.
Panel C: The genetic association and corresponding 95% confidence interval (CI) for each SNP (n = 696) with depression (x-axis) and Long COVID (y-axis) are plotted. The plot represents estimates from three methods: random-effects inverse weighted variance method, weighted median, and random-effects MR Egger.
The MR Egger intercepts are provided in **Supplementary Table 9a**.

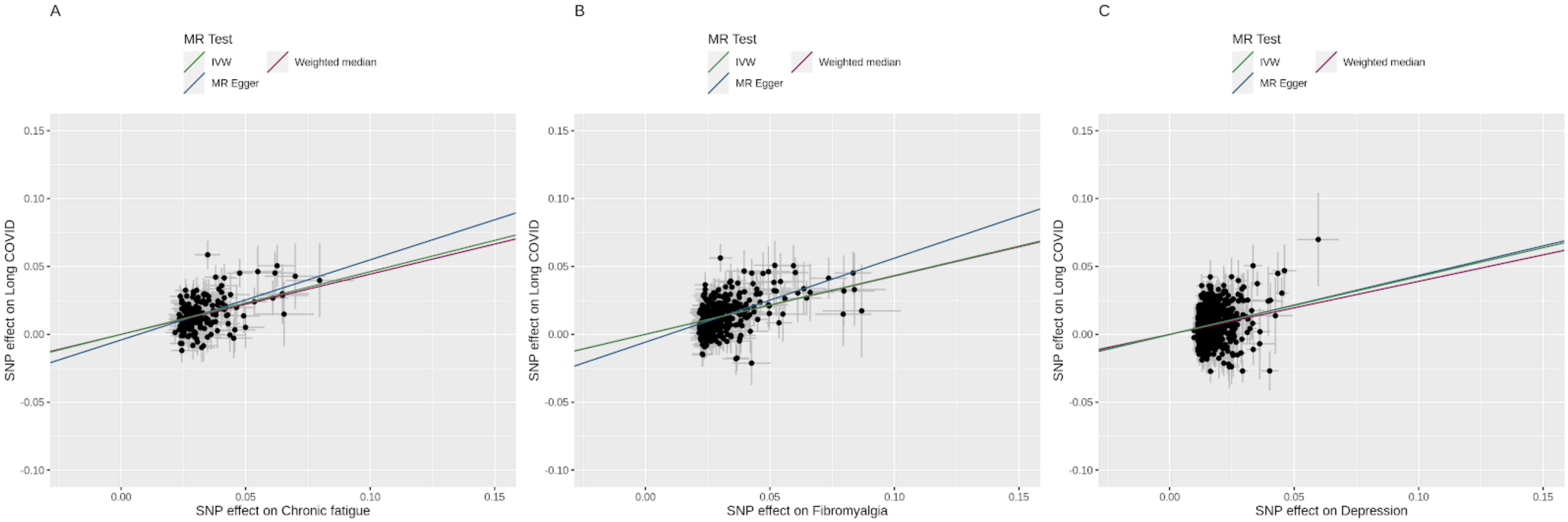

**Supplementary Figure 10: Forest plot representing genetically predicted effects of chronic phenotypes on Long COVID consortium using Mendelian randomization**

The estimates in the plot depict odds ratio and 95% confidence intervals. The summary statistics for Long COVID were obtained from COVID-19 Host Genetics Initiative.  Panel A represents MR estimates for chronic fatigue. The genetic instrument for chronic fatigue is derived using information from 186 SNPs (meanF statistic = 43.17). Panel B represents MR estimates for fibromyalgia. The genetic instrument for chronic fatigue is derived using information from 349 SNPs (meanF statistic = 45.04). Panel C represents MR estimates for depression. The genetic instrument for depression is derived using information from 696 SNPs (meanF statistic = 53.8). IVW = Inverse variance weighted.

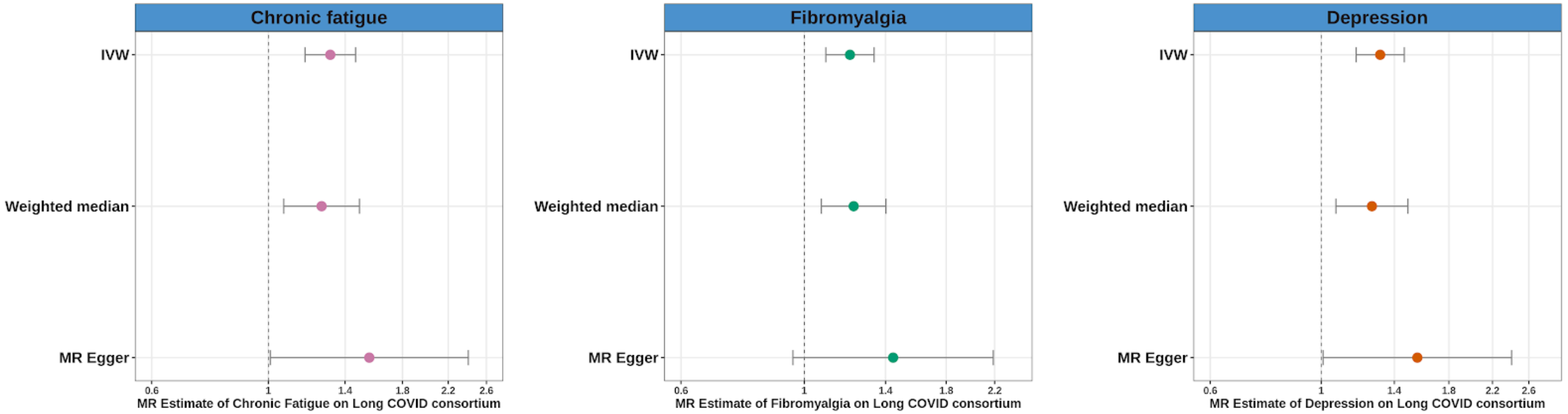

**Supplementary Figure 11: Forest plot representing genetically predicted effects of chronic phenotypes on COVID-19+ hospitalization from Long COVID consortium data using Mendelian randomization**
The estimates in the plot depict odds ratio and 95% confidence intervals. The summary statistics for COVID-19+ hospitalization were obtained from COVID-19 Host Genetics Initiative.  Panel A represents MR estimates for chronic fatigue. The genetic instrument for chronic fatigue is derived using information from 186 SNPs (meanF statistic = 43.17). Panel B represents MR estimates for fibromyalgia. The genetic instrument for chronic fatigue is derived using information from 349 SNPs (meanF statistic = 45.04). Panel C represents MR estimates for depression. The genetic instrument for depression is derived using information from 696 SNPs (meanF statistic = 53.8). IVW = Inverse variance weighted.

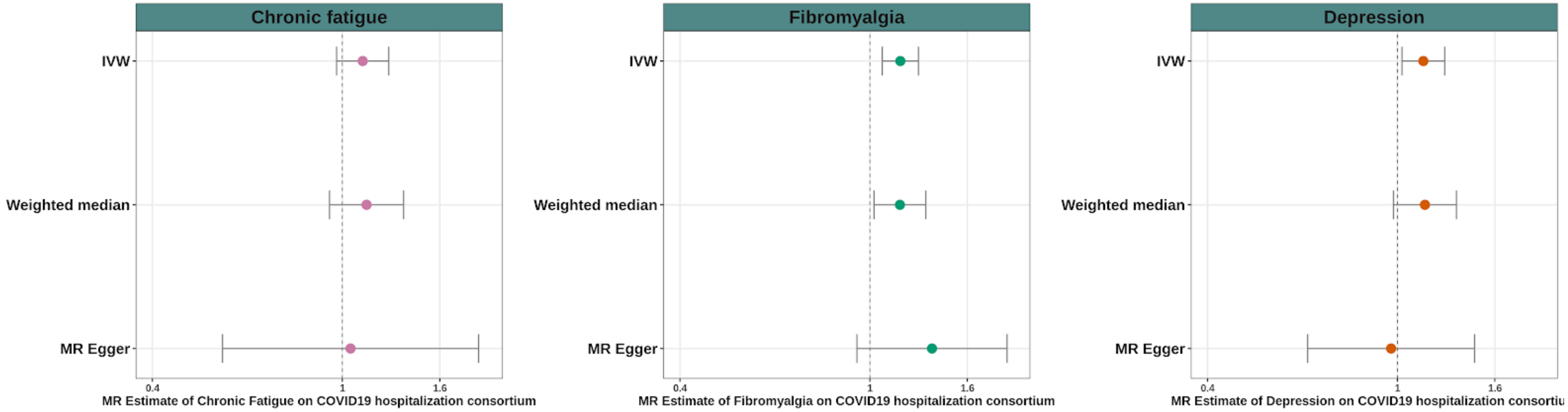

**Supplementary Figure 12: MR scatterplot of genetic associations between chronic phenotypes and Long COVID impact**

Panel A**:** The genetic association and corresponding 95% confidence interval (CI) for each SNP (n = 186) with chronic fatigue (x-axis) and Long COVID impact (y-axis) are plotted. The plot represents estimates from three methods: random-effects inverse weighted variance method, weighted median, and random-effects MR Egger.

Panel B**:** The genetic association and corresponding 95% confidence interval (CI) for each SNP (n = 349) with fibromyalgia (x-axis) and Long COVID impact (y-axis) are plotted. The plot represents estimates from three methods: random-effects inverse weighted variance method, weighted median, and random-effects MR Egger.

Panel C: The genetic association and corresponding 95% confidence interval (CI) for each SNP (n = 696) with depression (x-axis) and Long COVID impact (y-axis) are plotted. The plot represents estimates from three methods: random-effects inverse weighted variance method, weighted median, and random-effects MR Egger.

The MR Egger intercepts are provided in **Supplementary Table 9b**.

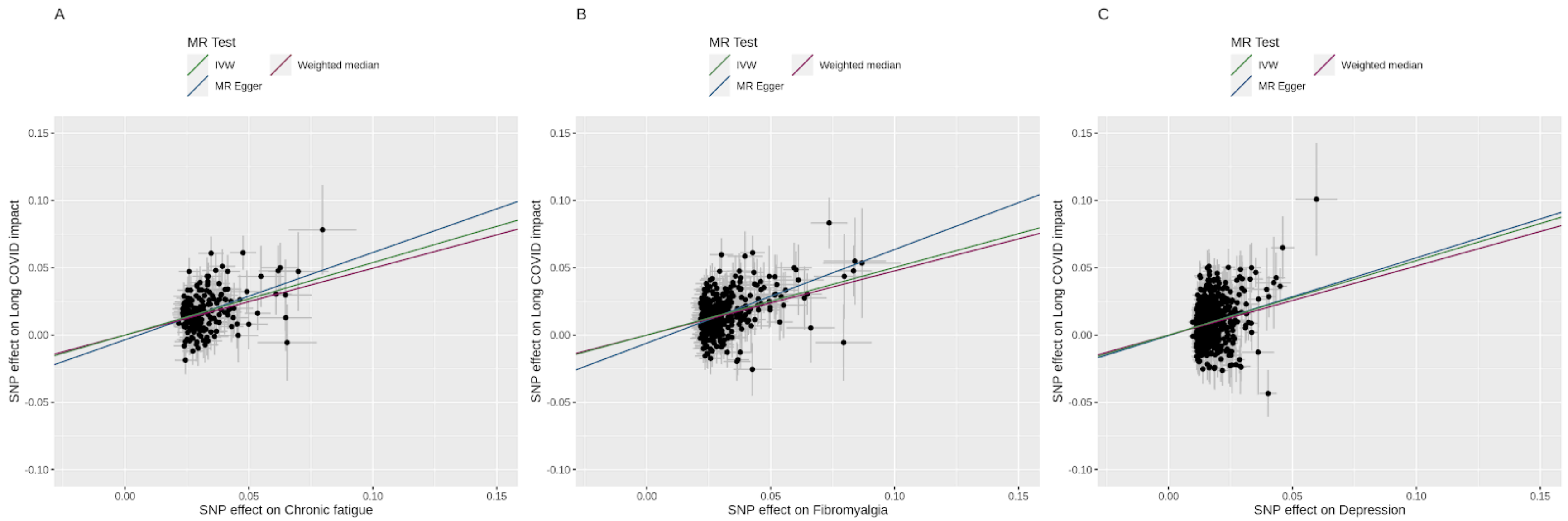

**Supplementary Figure 13: MR scatterplot of genetic associations between chronic phenotypes and COVID-19 hospitalization**

Panel A**:** The genetic association and corresponding 95% confidence interval (CI) for each SNP (n = 186) with chronic fatigue (x-axis) and COVID-19 hospitalization (y-axis) are plotted. The plot represents estimates from three methods: random-effects inverse weighted variance method, weighted median, and random-effects MR Egger.

Panel B**:** The genetic association and corresponding 95% confidence interval (CI) for each SNP (n = 349) with fibromyalgia (x-axis) and COVID-19 hospitalization (y-axis) are plotted. The plot represents estimates from three methods: random-effects inverse weighted variance method, weighted median, and random-effects MR Egger.

Panel C: The genetic association and corresponding 95% confidence interval (CI) for each SNP (n = 696) with depression (x-axis) and COVID-19 hospitalization (y-axis) are plotted. The plot represents estimates from three methods: random-effects inverse weighted variance method, weighted median, and random-effects MR Egger.

The MR Egger intercepts are provided in **Supplementary Table 9c**.

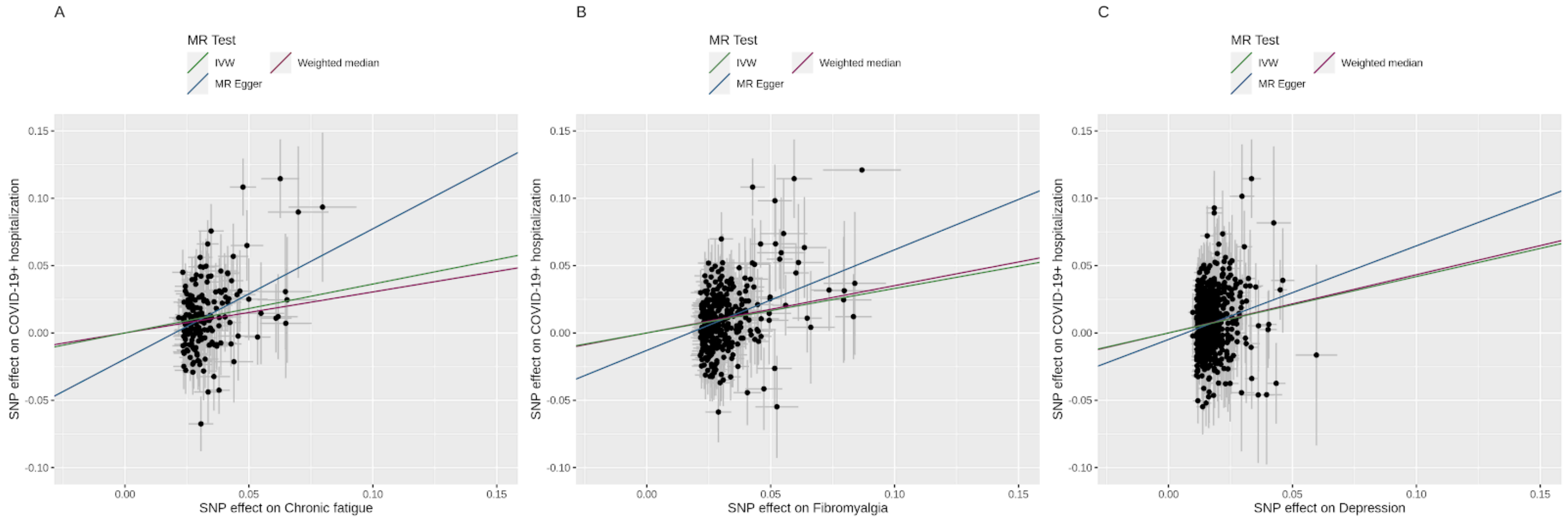

**Supplementary Figure 14**: **Forest plot representing genetically predicted effects of depression derived from external instruments on Long COVID using Mendelian randomization**

The estimates in the plot depict beta estimates and 95% confidence intervals. The summary statistics for depression were obtained from two sources. The genetic instrument [denoted by Meta-analysis in the figure] from meta-analysis from three largest multiple studies (UK BioBank, 23andMe study, and Psychiatric Genomics Consortium) included information from 87 SNPs (meanF statistic = 77.67). The second instrument [denoted by UK Biobank in the figure] derived from summary statistics of UK Biobank only included information from 35 SNPs (meanF statistic = 40.83). IVW = Inverse variance weighted. For comparison purposes, the estimates from the main MR analysis of depression on Long COVID using 23andMe data (**Figure3**) are included under sub-heading 23andMe

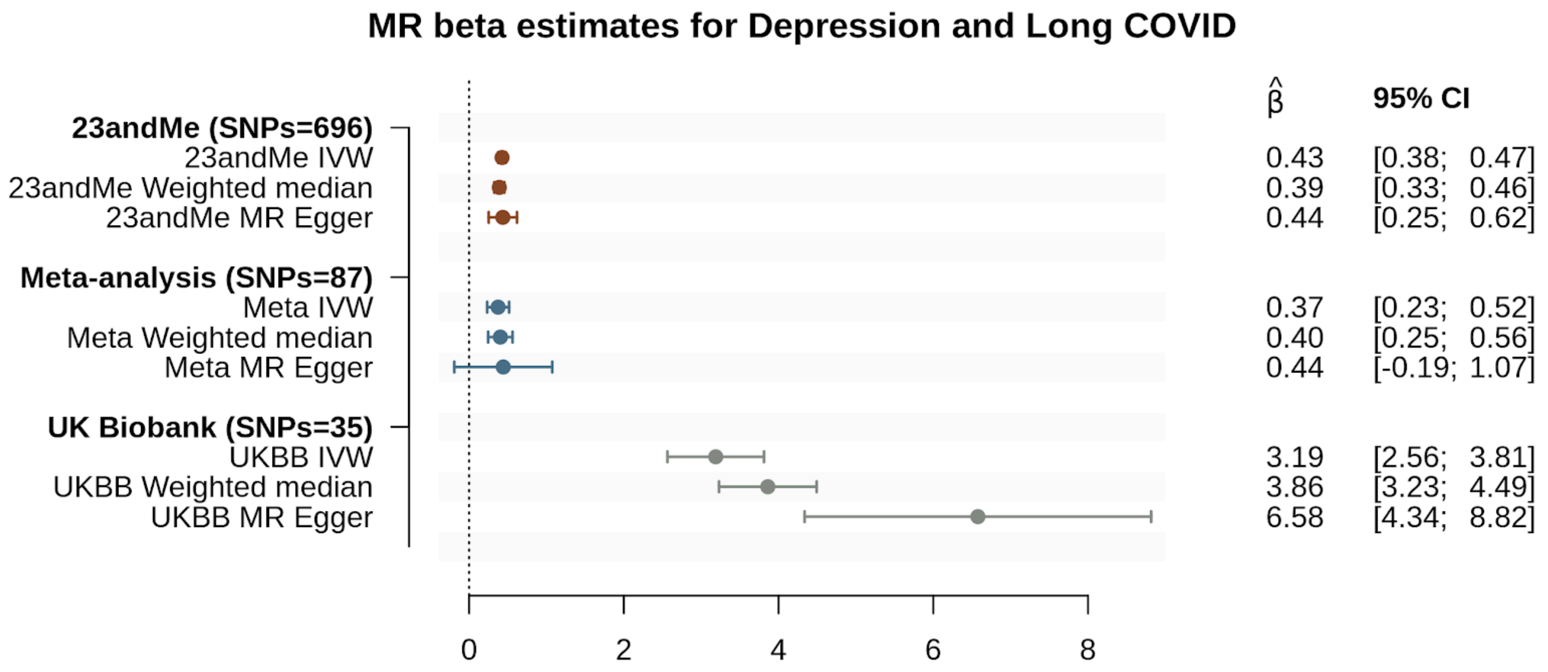
